# Prediction, prognosis and monitoring of neurodegeneration at biobank-scale via machine learning and imaging

**DOI:** 10.1101/2024.10.27.24316215

**Authors:** Anant Dadu, Michael Ta, Nicholas J Tustison, Ali Daneshmand, Ken Marek, Andrew B Singleton, Roy H Campbell, Mike A Nalls, Hirotaka Iwaki, Brian Avants, Faraz Faghri

**Affiliations:** Department of Computer Science, University of Illinois at Urbana-Champaign, Champaign, IL, 61820, USA; Center for Alzheimer’s and Related Dementias, National Institutes of Health, Bethesda, MD, 20892, USA; DataTecnica, Washington, DC, 20812, USA; University of Virginia, Dept of Radiology and Medical Imaging, Charlottesville, VA, 22903, USA; Department of Neurology, Boston Medical Center, Boston University School of Medicine, Boston, MA, 02118, USA; Laboratory of Neurogenetics, National Institute on Aging, National Institutes of Health, Bethesda, MD, 20892, USA; InviCRO LLC, Boston, Massachusetts

## Abstract

**Background:** Alzheimer’s disease and related dementias (ADRD) and Parkinson’s disease (PD) are the most common neurodegenerative conditions. These central nervous system disorders impact both the structure and function of the brain and may lead to imaging changes that precede symptoms. Patients with ADRD or PD have long asymptomatic phases that exhibit significant heterogeneity. Hence, quantitative measures that can provide early disease indicators are necessary to improve patient stratification, clinical care, and clinical trial design. This work uses machine learning techniques to derive such a quantitative marker from T1-weighted (T1w) brain Magnetic resonance imaging (MRI).

**Methods:** In this retrospective study, we developed machine learning (ML) based disease-specific scores based on T1w brain MRI utilizing Parkinson’s Disease Progression Marker Initiative (PPMI) and Alzheimer’s Disease Neuroimaging Initiative (ADNI) cohorts. We evaluated the potential of ML-based scores for early diagnosis, prognosis, and monitoring of ADRD and PD in an independent large-scale population-based longitudinal cohort, UK Biobank.

**Findings:** 1,826 dementia images from 731 participants, 3,161 healthy control images from 925 participants from the ADNI cohort, 684 PD images from 319 participants, and 232 healthy control images from 145 participants from the PPMI cohort were used to train machine learning models. The classification performance is 0.94 [95% CI: 0.93-0.96] area under the ROC Curve (AUC) for ADRD detection and 0.63 [95% CI: 0.57-0.71] for PD detection using 790 extracted structural brain features. The most predictive regions include the hippocampus and temporal brain regions in ADRD and the substantia nigra in PD. The normalized ML model’s probabilistic output (ADRD and PD imaging scores) was evaluated on 42,835 participants with imaging data from the UK Biobank. There are 66 cases for ADRD and 40 PD cases whose T1 brain MRI is available during pre-diagnostic phases. For diagnosis occurrence events within 5 years, the integrated survival model achieves a time-dependent AUC of 0.86 [95% CI: 0.80-0.92] for dementia and 0.89 [95% CI: 0.85-0.94] for PD. ADRD imaging score is strongly associated with dementia-free survival (hazard ratio (HR) 1.76 [95% CI: 1.50-2.05] per S.D. of imaging score), and PD imaging score shows association with PD-free survival (hazard ratio 2.33 [95% CI: 1.55-3.50]) in our integrated model. HR and prevalence increased stepwise over imaging score quartiles for PD, demonstrating heterogeneity. As a proxy for diagnosis, we validated AD/PD polygenic risk scores of 42,835 subjects against the imaging scores, showing a highly significant association after adjusting for covariates. In both the PPMI and ADNI cohorts, the scores are associated with clinical assessments, including the Mini-Mental State Examination (MMSE), Alzheimer’s Disease Assessment Scale-cognitive subscale (ADAS-Cog), and pathological markers, which include amyloid and tau. Finally, imaging scores are associated with polygenic risk scores for multiple diseases. Our results suggest that we can use imaging scores to assess the genetic architecture of such disorders in the future.

**Interpretation:** Our study demonstrates the use of quantitative markers generated using machine learning techniques for ADRD and PD. We show that disease probability scores obtained from brain structural features are useful for early detection, prognosis prediction, and monitoring disease progression. To facilitate community engagement and external tests of model utility, an interactive app to explore summary level data from this study and dive into external data can be found here https://ndds-brainimaging-ml.streamlit.app. As far as we know, this is the first publicly available cloud-based MRI prediction application.

**Funding:** US National Institute on Aging, and US National Institutes of Health.

**Research in context:** *Evidence before this study:* We searched PubMed for articles published in English from database inception to May 11, 2023, about the use of machine learning on brain imaging data for Alzheimer’s disease (AD), dementia, and Parkinson’s disease (PD) populations. We used search terms “machine learning” AND “brain imaging” AND “neurodegenerative disorders” AND “quantitative biomarkers”. The search identified 25 studies. Most of these studies are focused on Alzheimer’s disease. They use machine learning to predict conversion from mild cognitive impairment to dementia or to build a classification tool. Many studies also focused on positron emission tomography (PET) images rather than cost-effective T1w MRI images in their analysis. None of the studies have focused on detecting disease during the asymptomatic phase of dementia and PD. Identified studies are limited in sample size (order of hundred samples) and extracted features. The assessments of the clinical utility of machine learning models’ predicted disease probabilities are scarce. Significantly, no attempts were made to validate the algorithm in an external cohort. In this work, we have limited our review to scientific studies that are transparent and reproducible, including those that provide code and validate their findings on a reasonable sample size.

*Added value of this study:* This study developed machine learning based quantitative scores to measure the risk, severity, and prognosis of Alzheimer’s disease and related dementias (ADRD) and Parkinson’s disease (PD) using brain imaging data. Neurodegenerative disorders affect multiple body functions and exhibit significant etiology and clinical presentation variation. Patients with these conditions may experience prolonged asymptomatic periods. Disease-modifying therapies are most effective during the early asymptomatic stage of the disease, making early intervention a crucial factor. However, the lack of biomarkers for early diagnosis and disease progression monitoring remains a significant obstacle to achieving this goal. We leveraged disease-specific cohorts ADNI (1,826 images from 731 dementia participants) and PPMI (684 images from 329 PD participants) to develop a machine learning classifier for AD and PD detection using T1w brain imaging data. We obtain disease-specific imaging scores from these trained models using the normalized disease probability score. In a sizable external biobank, UK Biobank (42,835 participants), we found these scores show strong predictive power in determining the occurrence of PD or dementia during a 5-year followup. The occurrence of PD increased stepwise over ascending imaging score quantiles representing heterogeneity within the PD population. Imaging scores are also associated with pathological and clinical assessment measures. Our study indicates this could be a single numeric indicator representing disease-specific abnormality in T1w brain imaging modality. The association of imaging scores with the polygenic risk score of related disorders implies the genetic basis of these scores. We also identified top brain regions associated with dementia and Parkinson’s disease using feature interpretation tools.

*Implications of all the available evidence:* The findings should improve our ability to create practical passive surveillance plans for individuals with a heightened risk of occurrence of neurodegenerative disease. We have shown that imaging scores complement other risk factors, such as age and polygenic risk scores for early detection. The integrated model could serve as a tool for early interventions and study enrollment. Understanding the genetic basis of imaging scores can provide valuable insights into the biology of neurodegenerative disorders. Additionally, these high-accuracy models able to facilitate accurate early detection at the biobank scale can empower precision medicine trial recruitment strategies as well as paths of care for the future. We have included the development of an interactive web server (https://ndds-brainimaging-ml.streamlit.app) that empowers the community to process their own data based on our models and explore the utility and applicability of these findings for themselves. Users can easily upload a Nifti or DICOM file containing their MRI image, and we handle the entire pre-processing and prediction process. All computations are performed on the Google Cloud Platform. In addition, we provide an interpretation of the ML prediction highlighting areas of the brain that have contributed to the decision and a what-if-analysis tool where users explore different scenarios and their effect on prediction.

## Introduction

Neurodegeneration refers to the progressive loss of neuronal function or of neurons themselves. Alzheimer’s disease and related dementias (ADRD) and Parkinson’s disease (PD) are the most common forms of neurodegeneration affecting millions of people worldwide (Scheltens et al. 2016; Bloem, Okun, and Klein 2021; Holtzman, Morris, and Goate 2011). ADRD is predominantly characterized as a cognitive or behavioral disorder, while PD mainly affects motor skills (Gan et al. 2018). However, these are multisystem disorders affecting both the brain and the body. Further, both diseases exhibit substantial phenotypic heterogeneity with clinical manifestations varying by educational attainment, onset age, progression rate, or the constellation of motor/non-motor features (Fereshtehnejad et al. 2017; Satone et al. 2020; Dadu et al. 2022; Murray et al. 2011; Lam et al. 2013). ADRD and PD are now recognized as forming a phenotypic spectrum rather than discrete categories. Therefore, binary assignment cannot capture the complexity of ADRD and PD. Hence, probabilistic indicators of disease are required. Such quantitative markers can also function as surrogate endpoints to increase clinical trials’ ability to monitor treatment effects (Fleming and DeMets 1996).

Clinical diagnosis of neurodegenerative disease is preceded by a potentially long asymptomatic phase (Katsuno et al. 2018). Identifying the disease during patients’ pre-symptomatic phase may improve the success rate of disease-modifying therapies (Rasmussen and Langerman 2019). Abnormal anatomical changes may occur during the pre-symptomatic phase of ADRD and PD (Jack and Holtzman 2013). Further, the risk of getting these disorders highly depends on heritable factors, with more than 75 Alzheimer’s disease (AD) and 90 PD-associated genetic risk loci already identified (Bellenguez et al. 2022; Nalls et al. 2019). Together, genetic and imaging data modalities – which are longitudinally sensitive – are complementary predictors of clinical outcomes during the asymptomatic period of neurodegeneration.

Machine learning approaches have facilitated the analysis of a large number of brain structural features from T1-weighted (T1w) Magnetic resonance imaging (MRI) (Pellegrini et al. 2018). Machine learning models have shown success in accurately diagnosing AD (post-clinical diagnosis) (Pan et al. 2020; Huang et al. 2019), as well as predicting the conversion from mild cognitive impairment to AD (pre-clinical diagnosis) (Eskildsen et al. 2013; Spasov et al. 2019, Popuri et al, 2020). These approaches have been primarily used as a classification tool to discriminate patients from healthy individuals (binary classification) rather than quantifying disease continuously. Further, very few studies have validated their findings in an independent cohort (Bzdok, Varoquaux, and Steyerberg 2021). While T1-weighted MRI is commonly used to study AD, its application in PD is limited due to the need for more fine-grained features to analyze neuronal loss in the substantia nigra, the area most affected in PD (Schwarz et al. 2011, Popuri et al, 2020). We illustrated the utility of MRI imaging as a complementary measure for composite multi-modal biomarker panels, alongside other risk factors such as age, sex, and polygenic risk score, enhancing the performance of the survival model. Moreover, these studies are limited in the number of training samples and extracted features from MRI images. Finally, none of the studies have a web application for early detection of ADRD or PD using MRI, which stands out as another key differentiator in our work.

In the following, we present how we developed quantitative markers of ADRD and PD from brain imaging data using machine learning techniques and investigated their association with clinical outcomes during the pre-diagnosis and post-diagnosis phases. We evaluated the contribution of imaging scores, in combination with genetic risk factors to predict the likelihood of developing the disease later in life. In the post-diagnosis phase, we assessed the potential of imaging scores as a disease monitoring marker by examining their association with clinical assessments and relevant pathological biomarkers. We leverage disease-specific cohorts to train the machine learning models and evaluate machine learning-generated imaging scores in a large external biobank. The cost-effective biomarker using T1 brain imaging could pave the way for passive surveillance of the healthcare system for neurodegenerative disorders at a biobank scale. **Figure 1** highlights the workflow for our analysis.

**Figure 1:**
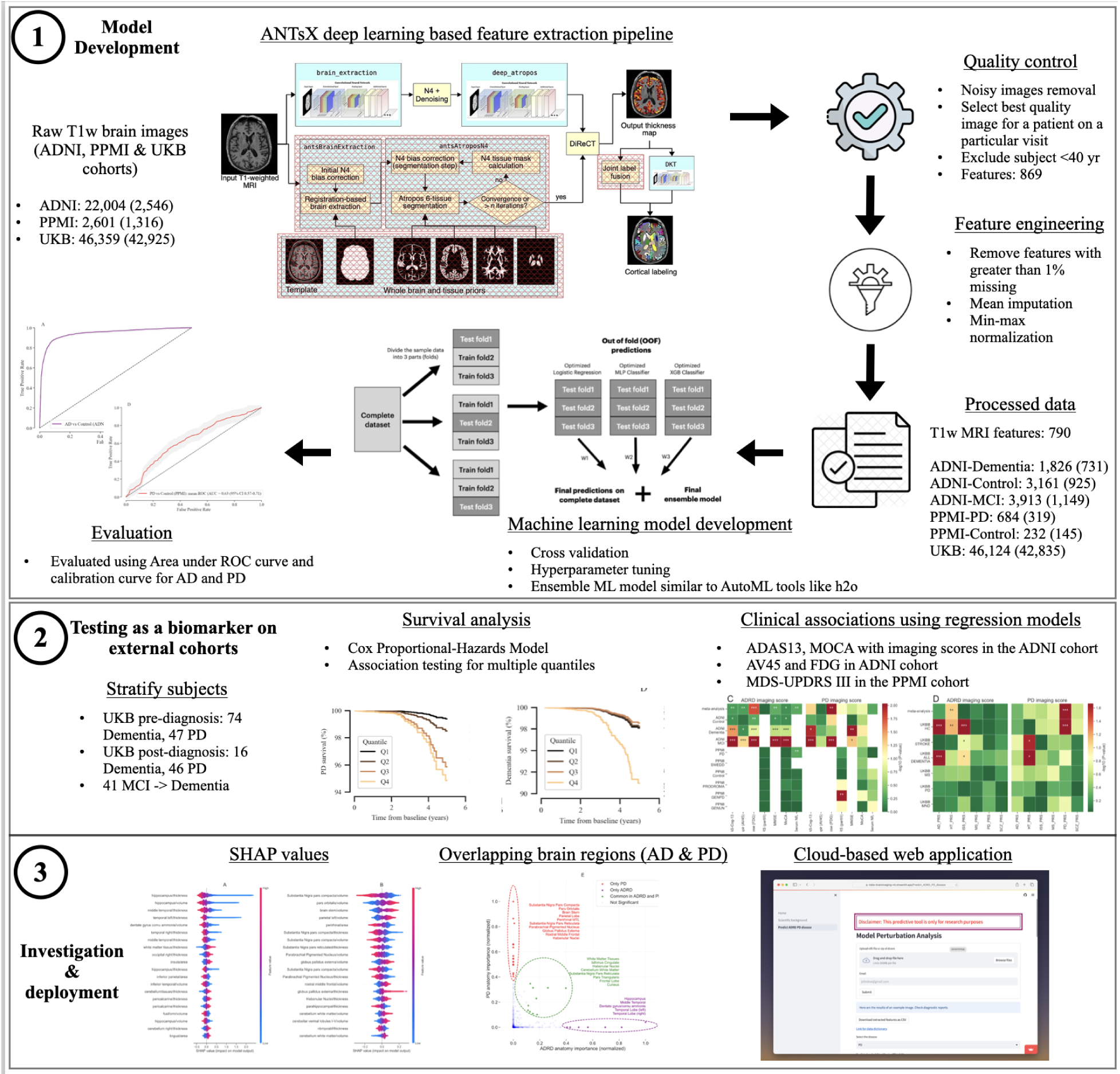
Workflow of analysis and model development.

## Methods

### Study design

We trained a supervised classification model using brain structural features extracted from T1w MRI brain images in PD-specific and AD-specific cohorts. Normalized disease probability scores were obtained after inference with a trained machine learning model. These scores were evaluated as a quantitative ADRD and PD disease marker in a large external biobank.

We trained and internally validated the AD classification model using the T1w MRI data from the Alzheimer’s Disease Neuroimaging Initiative (ADNI, https://adni.loni.usc.edu) cohort, which also allowed for examining the association of imaging scores with AD biomarkers and assessment tests. It consists of 731 dementia cases and 925 controls. Similarly for PD, Parkinson’s Progression Marker Initiative (PPMI, http://www.ppmi-info.org) was used, which includes 319 PD cases, and 145 controls (Table 1). We assessed the association of imaging scores with relevant clinical outcomes in 42,835 (22,588 female, 20,247 male) participants from the UK Biobank (UKB, https://www.ukbiobank.ac.uk) with T1w brain MRI. All participants and their study partners provided their consent, accepting their engagement for the data collection. The study protocols for ADNI, PPMI, and UKB were approved by the Institutional Review Board. Access and use of data from the UK Biobank were approved under application number 33601.

**Table 1:** Characteristics of participants in the training and internal validation set.

|  | PPMI |  | ADNI |  |
| --- | --- | --- | --- | --- |
|  | Controls<br>(I=232, N=145) | PD<br>(I=684, N=319) | Controls<br>(I=3,161, N=925) | Dementia<br>(I=1,826, N=731) |
| Mean age at baseline (SD), years | 62 (10) | 62 (9) | 73 (7) | 76 (8) |
| Mean age at T1w MRI performed (SD), years | 62 (10) | 63 (9) | 75 (7) | 76 (8) |
| Sex<br>Female<br>Male | 47 (32%)<br>98 (68%) | 111 (35%)<br>208 (65%) | 457 (56%)<br>358 (44%) | 237 (42%)<br>334 (58%) |
| Race<br>White<br>Black or African American<br>Asian<br>Other / unknown | 136 (93%)<br>7 (5%)<br>1 (1%)<br>2 (1%) | 296 (93%)<br>5 (2%)<br>6 (2%)<br>12 (3%) | 803 (87%)<br>75 (8%)<br>26 (3%)<br>24 (2%) | 683 (93%)<br>26 (4%)<br>16 (2%)<br>6 (1%) |
| Baseline clinical outcomes<br>MOCA<br>MMSE<br>MDS-UPDRS III<br>ADAS-Cog-13 | 28 (1)<br>NA<br>1 (2)<br>NA | 27 (2)<br>NA<br>22 (9)<br>NA | 26 (3)<br>29 (1)<br>NA<br>9 (4) | 17 (5)<br>23 (3)<br>NA<br>29 (8) |

ADNI, Alzheimer's Disease Neuroimaging Initiative; PPMI, Parkinson's Progression Markers Initiative; PD, Parkinson's disease; T1w MRI, T1 weighted image magnetic resonance imaging; MOCA, Montreal Cognitive Assessment; MMSE, Mini-Mental State Examination; MDS-UPDRS III, MDS-Unified Parkinson's Disease Rating Scale Part III; SD, standard deviation; I, number of images; N, number of individuals.

**Table 2:**
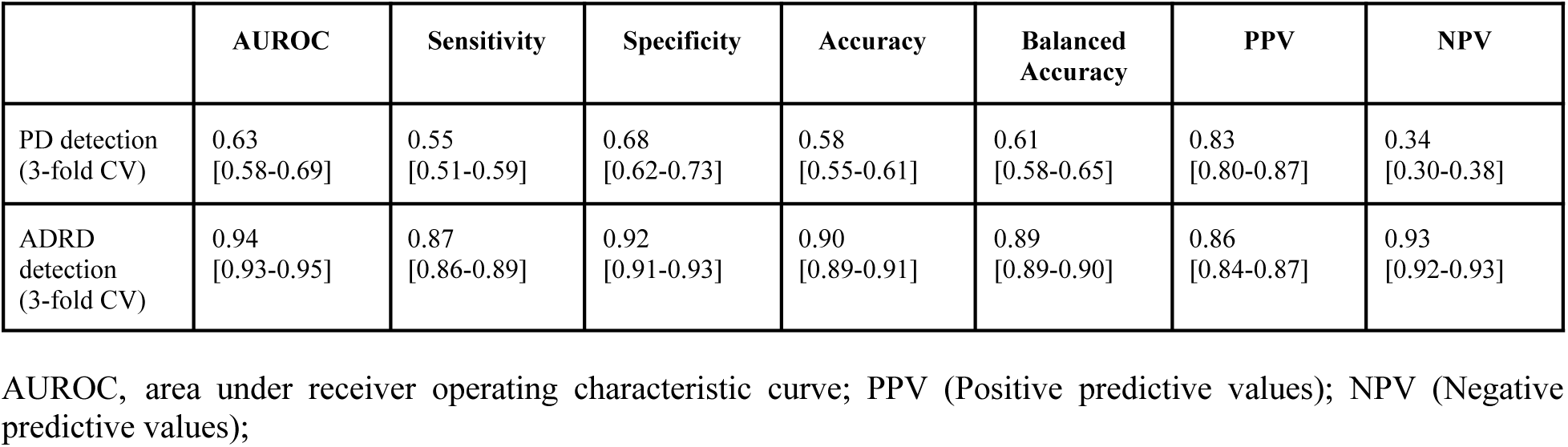
Classification performance of machine learning model for PD and AD detection.

### Study participants

The study included two disease-specific cohorts (ADNI, PPMI) and an external large biobank (UK Biobank). ADNI enrolls participants between the ages of 55 and 90 recruited at 57 sites in the United States and Canada. After obtaining informed consent, participants undergo a series of initial tests repeated at intervals over subsequent years, including a clinical evaluation, neuropsychological tests, genetic testing, lumbar puncture, and MRI and PET scans. PPMI Clinical protocol is designed to acquire comprehensive longitudinal within-participant data from approximately 4,000 participants enrolled at about 50 sites worldwide. The quantitative marker’s performance was assessed in the UK Biobank, a community-based cohort of over 500,000 individuals, mainly of British self-reported ethnicity, between the ages of 40 and 69, recruited from across the UK between 2006 and 2010. We only included participants with T1w MRI data available, and the patients were randomly selected for imaging by UK Biobank cohort setup, avoiding the possibility of selection bias. The details of the number of participants used in the analysis are shown in **Figure 1** and **Table 1**.

### ADRD and PD diagnosis

In the PPMI cohort, all PD patients fulfilled the UK Brain Bank Criteria, and healthy control subjects had no clinical signs suggestive of parkinsonism, no evidence of cognitive impairment, and no first-degree relative diagnosed with PD (Sudlow et al. 2015). For the ADNI cohort, the Alzheimer’s disease (AD) diagnosis is based on the criteria established by the National Institute on Aging and the Alzheimer’s Association (NIA-AA). The diagnosis of MCI was based on the core clinical criteria for MCI established by the NIA-AA workgroups (Jack et al. 2018), and the criteria were modified from the criteria proposed by Petersen et al. (Petersen et al. 2021). Since the accurate confirmation of dementia type uses the post-mortem brain, we are considering the AD cases in ADNI as ADRD rather than specific to AD. In the UK Biobank, the diagnosis date and the participants with all causes of dementia were identified using Field 42018. Field 42032 was used for PD (for a detailed description of Field in UKB, refer to https://biobank.ndph.ox.ac.uk/showcase/). The age distribution is shown in **Supplementary Figure 1**.

### Clinical features obtained from T1w brain imaging data

We use functionality from the Advanced Normalization Tools (ANTsX) Ecosystem (Tustison et al. 2021) to extract structural features such as the volume and area of different brain regions from T1w magnetic resonance images (MRI). This includes features from more nuanced and smaller brain regions, such as the brain stem and substantia nigra. These ANTsX-based features have demonstrated significantly improved performance in terms of age and gender prediction compared to its counterpart Freesurfer (Tustison et al. 2021). For our study, we used the ANTsPyT1w pipeline, available at https://github.com/stnava/ANTsPyT1w, to extract structural features from the T1w images yielding a total of 869 features per image. A list of all features with missing data details is shown in **Supplementary Table 1**. After filtering and excluding 79 features with more than 1% missing values, we trained the ML model using 790 continuous features. Missing values for variables with <1% missing data were imputed with the column mean. We performed min-max normalization to scale the range of all features between 0 and 1. Our study is the first to have a consistent pipeline for extracting such a large number of features from two large neurodegeneration-specific and biobank-scale datasets.

### Development of the machine learning model

We develop a stacked ensemble of three supervised machine learning algorithms (Neural network, XGBoost, Logistic regression) to train a classification model for Parkinson’s disease (PD) and Alzheimer’s disease (AD) in disease-specific cohorts. Stacked ensemble is a linear blend of multiple base models. These base models are optimized on average performance estimates from test folds (using cross-validation) through hyperparameter tuning. Once we obtain the optimized base models, the stacked ensemble model is developed based on linear blending on out-of-fold predictions. This approach helps avoid overfitting and provides better estimates of model performance, as it utilizes all the available data. Supplementary Figure 2 shows the schematic view of the model development procedure. After extensive hyperparameter tuning, the best performing ensemble model was trained and selected. Feature selection was performed using the Least Absolute Shrinkage and Selection Operator (LASSO) (Tibshirani 1996) as a part of the hyperparameter tuning procedure. The effectiveness of our classifier was validated using a 3-fold cross-validation approach. We selected 3-fold cross-validation to balance computational efficiency and validation accuracy. While higher fold cross-validation can yield more stable performance estimates, it demands significantly more time and computational resources, particularly when using multiple algorithms and extensive hyperparameter tuning. Model performance was evaluated based on various metrics, including accuracy, area under the receiver operating characteristic curve (AUC), sensitivity, and specificity. Although longitudinal data was used for training the model, we ensured that the cross-validation folds were split so that the same individual did not appear in both the training and test folds. We did not employ nested approaches or multilevel models to further account for clustering at the participant level. Since we are focusing on generating disease severity scores and incorporating all the images is a better choice. This approach ensured that the model performance reports have little bias with overfitting to diagnostic labels. The 95% confidence intervals (CIs) on diagnostic and prognostic performance estimates were calculated using a percentile bootstrap with 1000 samples. We employed the Shapley Additive Explanations (SHAP) approach to assess the impact of each feature on the machine learning model predictions (Lundberg and Lee 2017). SHAP values are derived from game theory and approximate a feature’s effect on the model. SHAP enhances understanding by creating accurate explanations for each observation. The interactive website (https://ndds-brainimaging-ml.streamlit.app) was developed as an open-access and cloud-based platform for researchers to investigate the top features of the machine learning models developed and how these may influence the disease probability scores (refer to **Feature importance and website development section in Supplementary Material**). Finally, the probability scores were normalized using log-odds transformation to generate the disease-specific quantitative markers. End-to-end deep learning pipelines might be useful to mine complete imaging information. However, it is important to note that deep learning requires a significant amount of training data and remains a ‘black box’. Since we are focusing more on clinical applicability, we have limited our work to transparent and reproducible models. Using interpretable and reproducible deep learning models for brain imaging would be a promising future direction.

### Features from UK Biobank cohort

We computed ADRD and PD imaging scores for all participants in the UK Biobank who have available T1w brain MRI. In our analysis, we also considered known risk factors associated with neurodegenerative disorders comprising age at recruitment (Field 21022), Date of attending assessment center (Field 53), Sex (Field 31), Townsend deprivation index at recruitment (Field 22189), Standard polygenic risk score (PRS) for Alzheimer’s disease (Field 26206) and Standard PRS for Parkinson’s disease (Field 26260) available in UKB database. PRS was calculated based on the methodology described in (Thompson 2022).

### Clinical assessments and MCI convertors to Dementia

We evaluated the association of imaging scores with standard clinical assessments used for diagnosing dementia, such as MoCA score, MMSE, and ADAS-Cog, available in the ADNI database. Additionally, we utilized pathological biomarkers of AD, including AV45 and FDG, to validate the imaging scores. We also assessed the predictive value of imaging scores for the conversion from MCI to dementia. To validate with respect to PD-related clinical measurements, we extracted MDS-UPDRS-III, MoCA, and SerumNfl biomarkers from the PPMI database.

### Statistical analysis

We employed a Cox proportional hazards regression model to examine the correlation between imaging scores and the clinical diagnosis time of participants who had imaging data before diagnosis. These models were adjusted for relevant covariates to calculate hazard ratios (HR) for imaging scores. We censored events for individuals with an attained survival of greater than five years from the baseline (time at which participant enters the study) because of insufficient numbers of PD or Dementia events beyond that. To estimate survival curves, we used imaging score quantiles. We reported the hazard ratio, p-values, and 95% CI. The overall performance was measured using the concordance index, while the time-dependent AUC metric was used to evaluate the model’s performance in terms of stratification based on the time from the event (refer to the **Evaluation metrics** section in **Supplementary Material**). Linear regression was used to associate the polygenic risk score with the imaging score and to determine the relation between imaging scores and disease monitoring clinical assessment tests (refer to **Association testing** section in **Supplementary Material**). All statistical tests and plots were created using Python (version 3.8). We utilized the PPMI and ADNI cohorts to associate clinical and pathological biomarkers, and the UK Biobank to associate with polygenic risk scores.For the survival analysis model, we used Time-dependent AUC and C-index, which are described in the Evaluation Metrics section of the Supplementary Material.

## Results

Feature extraction using AntsPyT1w from T1w MRI images was completed for 2,546 participants (22,004 images) in the ADNI dataset, 1,316 participants (2,601 images) in the PPMI cohort, and 42,925 participants (46,124 images) available in the UK Biobank cohort. Each image has an automated quality score from the ANTsX pipeline that is used to remove noisy images. There may be multiple images for a single participant on a particular visit, so we chose the best quality images out of these. Further, we excluded participants who were less than 40 years old. We trained an ADRD classification model on 1,826 dementia and 3,161 control image samples from the ADNI cohort. We also trained a PD classification model on 684 dementia and 232 control image samples from the PPMI cohort (demographics shown in **Table 1**).

Our ensemble stack includes logistic regression, neural networks, and XGBoost. The details of the trained ensemble model are in **Supplementary Table 2**. For Alzheimer’s Disease prediction, the contributions are 44% from logistic regression, 50% from neural networks, and 6% from XGBoost. For Parkinson’s Disease prediction, logistic regression contributes 42%, neural networks 55%, and XGBoost 3%. We correctly distinguish images with ADRD based on imaging features with an AUC of 0.94 (95% CI: 0.93-0.96) and PD with an AUC of 0.63 (0.56-0.70) at cross-validation (subject-based) (**Figure 2A**, **Figure 2D**). Both models are very well calibrated for the ADRD detection model and PD (**Figure 2B**, **Figure 2E**). The well-calibrated model is necessary for our study as we focus on the usability of probabilistic scores obtained from machine learning models. The distribution of disease probabilities and log-odds normalized probabilities on different cohorts is shown in **Supplementary Figure 3**. The quantification scores allow us to extend the model trained on the post-diagnosis phase to develop a model for early detection of AD and PD during the pre-diagnostic phase.

**Figure 2.**
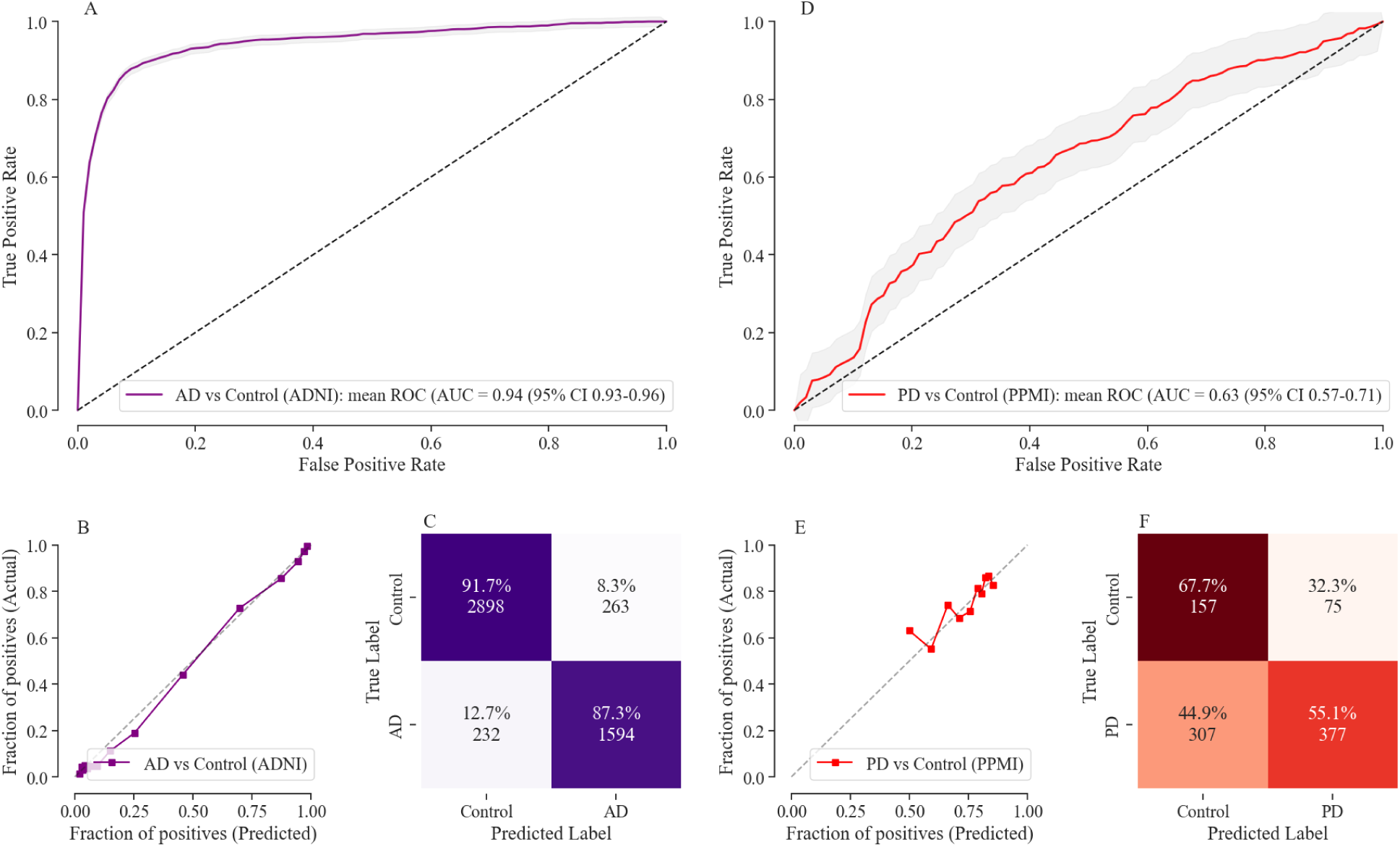
Performance of the machine learning model for the detection of AD in the ADNI and PD in the PPMI cohort following stratified 3-fold CV (based on individuals) evaluation scheme. **(A)**, **(D)** The machine learning model discriminated Dementia from healthy controls with cross-validated AUROCs of 0·94 (95% CI 0·93–0·96), and PD from healthy controls with AUROCs of 0.63 (95% CI 0·57–0·71). **(B)**, **(E)** Shows the calibration curves for trained classifiers, demonstrating that the model is well calibrated and not making overconfident or underconfident probabilistic predictions. **(C)**, **(F)** Shows the confusion matrix on cross-validation folds for AD and PD detection respectively at the threshold with maximum F1 score. For the AD classification model, sensitivity is 92.59%, specificity is 85.84%, negative predictive value (NPV) is 87.29%, precision is 91.68%, and accuracy is 90.07%. For the PD classification model, sensitivity is 33.84%, specificity is 83.41%, NPV is 55.12%, precision is 67.67%, and accuracy is 58.30%. CV, cross-validation; AD, Alzheimer’s Disease; PD, Parkinson’s Disease; ADNI, Alzheimer’s Disease Neuroimaging Initiative; PPMI, Parkinson’s Progression Markers Initiative; AUROC, area under the receiver operating characteristic curve.

We evaluated the association between ADRD/PD imaging score and the occurrence of all cause dementia and PD in the UK Biobank during the pre-diagnostic phase using survival analysis. In UKB, 66 individuals were diagnosed with dementia and 40 with PD within 5 years from baseline (**Supplementary Figure 4**). ADRD imaging score was associated with a quantitative increase in the risk of getting dementia, with an adjusted HR of 1.76 (95% CI 1.50–2.05; p < 0.0001) per S.D. increase in ADRD imaging score. Also, PD imaging score was associated with a quantitative increase in the risk of getting a PD diagnosis, with an adjusted HR of 2.33 (95% CI 1.55–3.50; p < 0.0001) per S.D. increase in PD imaging score. Prevalence and risk of getting clinical diagnosis increased stepwise over ascending imaging quartiles: two (0.2%) of 1247 participants who had follow-up in the bottom quartile (Q1), and 20 (1.9%) of 1247 in the top quartile (Q4; HR: 8.5 [1.9–37.5]; p < 0.0001; **Figure 3**). We observed a continuous phenotypic spectrum for PD in which the imaging score (abnormality in T1w image) varies across the PD population during the pre-diagnostic phase. For dementia, we observed more homogeneous patterns, as most subjects (in quartile 1) demonstrated abnormality in T1w during the pre-diagnosis stage. The contribution of imaging score is further useful in the early detection of prognosis from MCI to dementia with the hazard ratio (2.99 [2.31–3.86]; p < 0.0001) per S.D. increase in ADRD imaging score. To maximize clinical utility, it is essential to obtain combined risk estimates from imaging and genetics to decide against (low risk) or for (high risk) taking clinical action.

**Figure 3.**
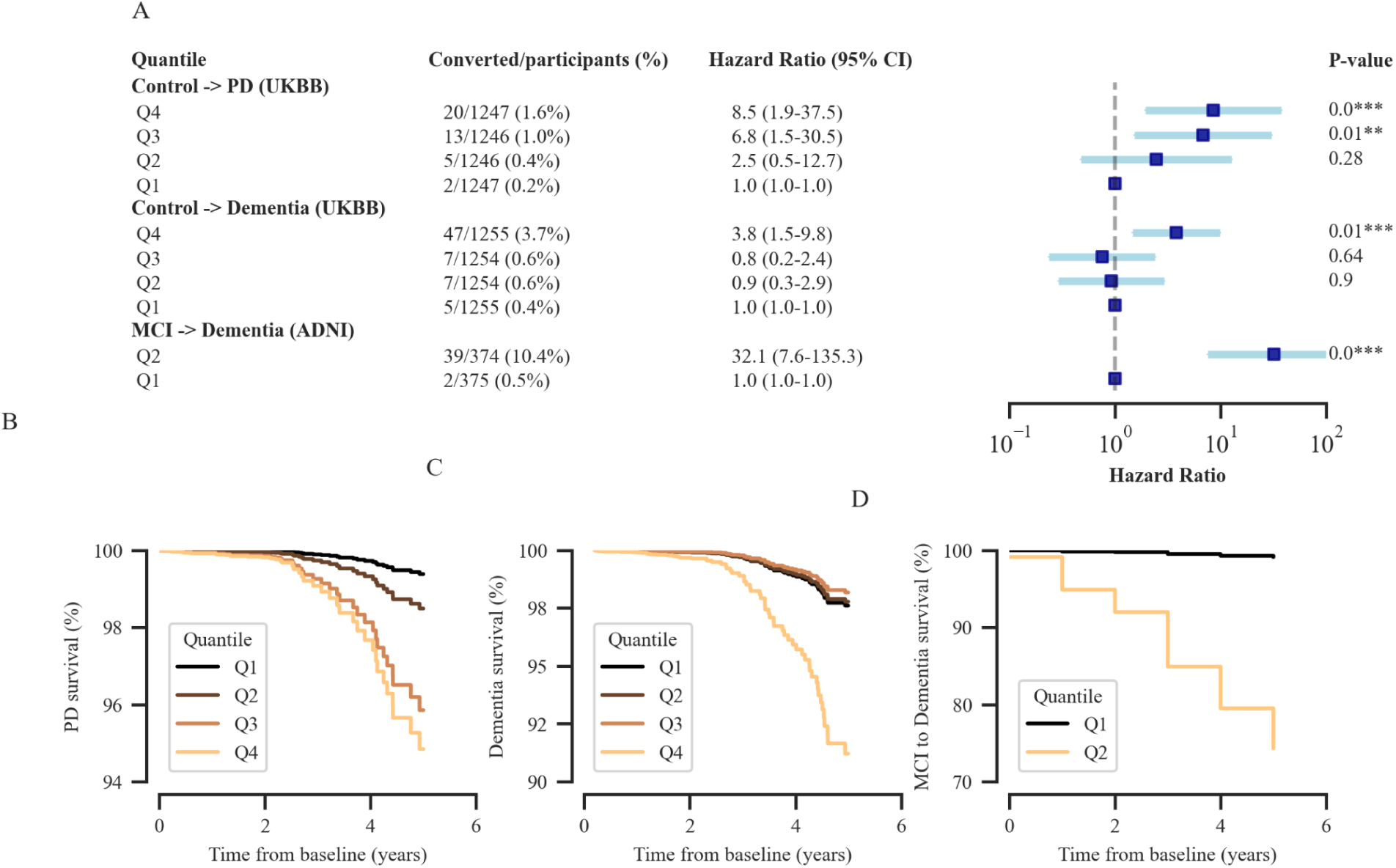
Testing utility of AD/PD imaging scores as a risk, prognostic, and monitoring biomarker. Imaging scores were stratified into multiple quantiles and adjusted HRs were compared with the lowest quantile. **(A)** Plot summarized the conversion from healthy control to PD (PD risk), healthy control to Dementia (Dementia risk) and MCI to Dementia (prognosis) with adjusted HR using Cox PH survival model. Both HR and converted percentage increased over ascending imaging score quantiles. The effect size is significantly different between the highest and lowest quantile for all the conversion as depicted in the forest plot. **(B)** Covariate-adjusted survival curves for patients for different quantiles with duration (in years) from the conversion to PD on the x-axis and fraction of individuals surviving free of conversion on the y-axis**, (C)** Covariate-adjusted survival curves for conversion to Dementia, and **(D)** Covariate-adjusted survival curves for conversion from MCI to Dementia. Higher imaging score quantiles had lower survival for conversion as compared with lower quantiles. HR, hazard ratio; Cox PH, Cox Proportional-Hazards.

We used time-dependent AUC and concordance index to characterize the discrimination potential of the imaging scores (**Table 3**). The integrated survival model (age, sex, Townsend deprivation index, polygenic scores, and imaging scores) achieves a time-dependent AUC of 0.86 [95% CI: 0.80-0.92] for dementia and 0.89 [95% CI: 0.85-0.94] for PD. The lower performance of PD detection in the PPMI cohort compared to the UKB cohort can be attributed to differences in the age distribution of PD diagnosis between the two cohorts (**Supplementary Figure 1, Supplementary Figure 5)**. We observed that the predictive significance of the imaging score in determining survival is more significant for subjects who converted within three years from baseline compared to those who converted after three years (relative improvement declines by 3.5% measured using time-dependent AUC). It is consistent for both dementia and PD conversion. It is a valuable result as it could guide the healthcare system to determine the duration between image collections.

**Table 3:** Performance of survival analysis algorithm using demographics, imaging score and polygenic risk scores.

|  | Imaging<br>HR<br>(95% CI) | Imaging<br>P-value | Concordance<br>Index<br>(95% CI) | Mean t-AUC<br>(95% CI) | Mean t-AUC<br>(95% CI)<br>< 3 years | Mean t-AUC<br>(95% CI)<br>≥ 3 years |
| --- | --- | --- | --- | --- | --- | --- |
| **PD risk prediction** |  |  |  |  |  |  |
| A + S + T + PD_PRS | NA | NA | 0.83<br>[0.78-0.89] | 0.87<br>[0.82-0.92] | 0.84<br>[0.77-0.93] | 0.91<br>[0.86-0.97] |
| PD_Imaging + A + S + T + PD_PRS | 2.33<br>[1.55-3.50] | 4.8e-5 | 0.86<br>[0.81-0.92] | 0.89 **<br>[0.85-0.94] | 0.87<br>[0.81-0.96] | 0.92<br>[0.89-0.99] |
| **AD risk prediction** |  |  |  |  |  |  |
| A + S + T + AD_PRS | NA | NA | 0.82<br>[0.76-0.89] | 0.84<br>[0.77-0.90] | 0.78<br>[0.66-0.92] | 0.89<br>[0.82-1.01] |
| ADRD_Imaging + A + S + T + AD_PRS | 1.76<br>[1.50-2.05] | 1.6e-12 | 0.86<br>[0.79-0.93] | 0.86 *<br>[0.80-0.92] | 0.81<br>[0.67-0.97] | 0.90<br>[0.84-1.01] |
| **MCI => Dementia risk prediction** |  |  |  |  |  |  |
| A + S + BV + AD_PRS | NA | NA | 0.52<br>[0.41-0.61] | 0.51<br>[0.37-0.63] | 0.55<br>[0.30-0.86] | 0.50<br>[0.33-0.64] |
| ADRD_Imaging + A + S + BV + AD_PRS | 2.99<br>[2.31-3.86] | 5.7e-17 | 0.73<br>[0.64-0.84] | 0.74 ***<br>[0.63-0.87] | 0.65<br>[0.40-0.97] | 0.73<br>[0.60-0.85] |

A, Age; S, Sex; T, Townsend; PD\_PRS, polygenic risk score of Parkinson's disease; PD\_Imaging, PD imaging score; AD\_PRS, polygenic risk score of Alzheimer's disease; ADRD\_Imaging, ADRD imaging score; BV, brain volume. Significance tests were performed using the DeLong comparison test within each category (\* indicates $P < 0.05$ , \*\* indicates $P < 0.001$ , and \*\*\* indicates $P < 0.0001$ ). For the conversion to mild cognitive impairment (MCI), the positive predictive value (PPV) is 0.12, and the negative predictive value (NPV) is 0.97, with a confusion matrix showing 528 true negatives (TN), 25 true positives (TP), 16 false negatives (FN), and 180 false positives (FP). In predicting Alzheimer's disease (AD), the PPV is 0.04, and the NPV is 0.99, with 3784 TN, 52 TP, 14 FN, and 1178 FP. For Parkinson's disease (PD) prediction, the PPV is 0.03, and the NPV is 0.99, with 2974 TN, 38 TP, 2 FN, and 1982 FP.

SHAP values provide in-depth analysis of machine learning classifiers by highlighting the top discriminating features of ADRD and PD. The biological plausibility of the models is supported by the feature importance plots, which highlight regions known to be seed regions for AD and PD pathology. In ADRD, features related to the hippocampus and temporal brain regions have the greatest discriminatory power (**Figure 4A**). Midbrain regions, including substantia nigra, brain stem, and parietal brain regions, help distinguish PD from controls (**Figure 4B**).

**Figure 4.**
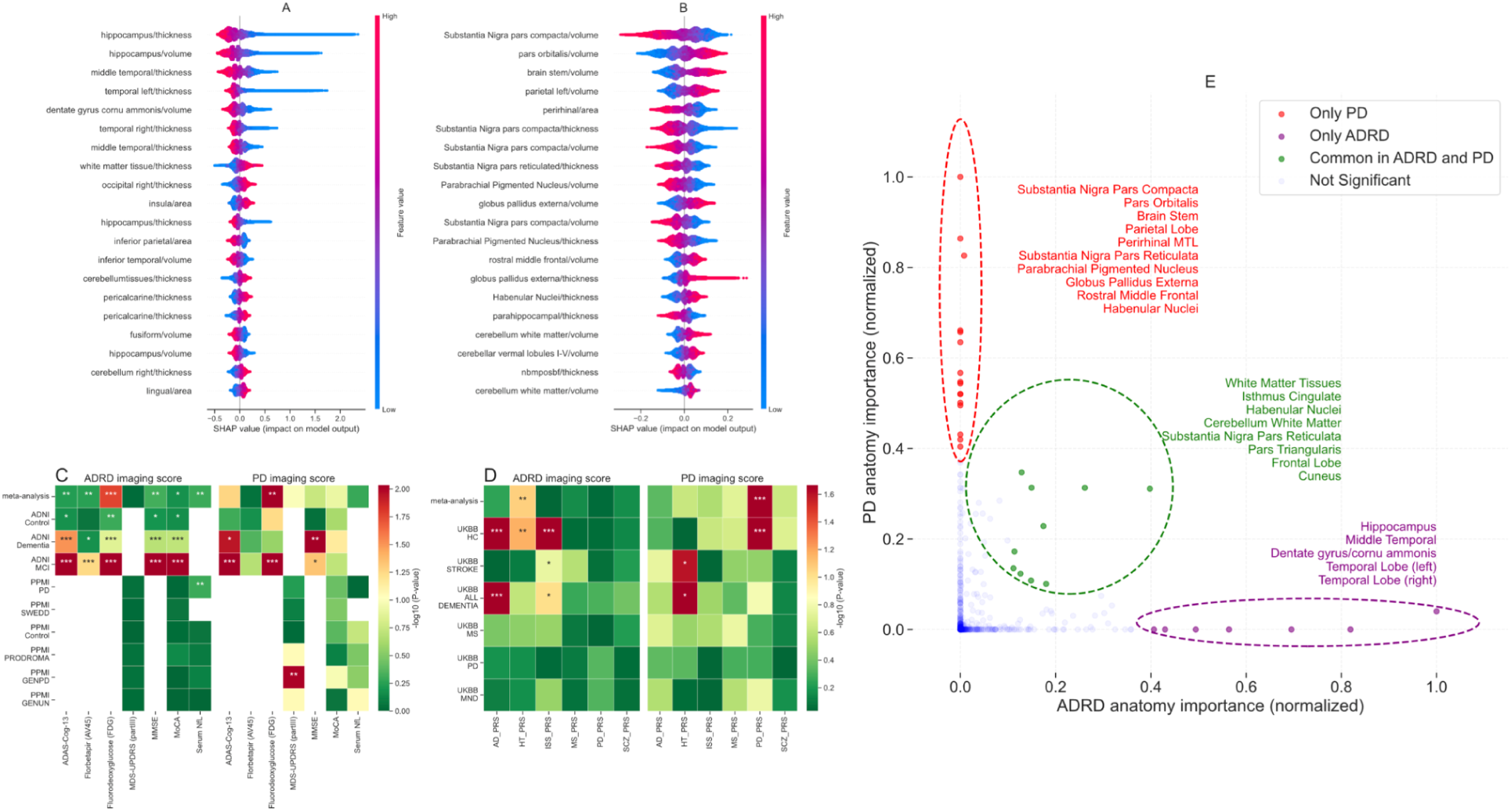
Interpretation of imaging scores and its association with polygenic risk scores. **(A)** Distribution of the top-20 features that had the most substantial effect on the ADRD imaging score (disease probabilities). Each point represents a patient and the amount of effect on model output for each feature depends on its SHAP value. For example, the effect of the hippocampus cortical thickness feature on model output is large and positive (high risk) when the patient has low values for hippocampus cortical thickness (more blue points are on the right side). Similarly, **(B)** shows the top features for PD imaging scores. **(C)** Shows the heatmap plot of P-values for the association testing between ADRD/PD imaging scores with clinical outcomes in ADNI and PPMI cohort stratified based on diagnosis status. **(D)** Heatmap plot showing association test of imaging scores with polygenic risk scores of related diseases in the UK biobank cohort. **(E)** Shows the primary anatomical regions associated with PD and ADRD, including regions that may overlap between these disorders. For more details of **(C)** and **(D)**, refer to the **Association testing** section in **Supplementary Material**. ADAS-Cog-13, Alzheimer’s Disease Assessment Scale-Cognitive Subscale; MoCA, Montreal Cognitive Assessment; MMSE, Mini-Mental State Examination; MDS-UPDRS III, MDS-Unified Parkinson’s Disease Rating Scale Part III; Serum Nfl, Serum neurofilament light; PPMI-SWEDD, Patients with scans without evidence of dopaminergic deficit in PPMI; PPMI-PRODROMAL, prodromal PD patients; PPMI-GENPD, Genetic affected PD; PPMI-GENUN, Genetic unaffected PD; AD_PRS, polygenic risk score of Alzheimer’s disease; HT_PRS, polygenic risk score of Hypertension; ISS_PRS, polygenic risk score of Ischemic stroke; MS_PRS, polygenic risk score of Multiple Sclerosis; PD_PRS, polygenic risk score of Parkinson’s disease; SCZ_PRS, polygenic risk score of Schizophrenia; UKBB-HC, healthy control in UK biobank (with no NDDs); UKBB-STROKE, Stroke patients in UK biobank; UKBB-ALL_DEMENTIA, all cause dementia patients in UK biobank; UKBB-MS, Multiple Sclerosis patients in UK biobank; UKBB-PD, Parkinson’s disease patients in UK biobank; UKBB-MND, Motor neurone disease patients in UK biobank; ***, P<0.001; **, P<0.005; *, P<0.01.

To assess the potential of imaging scores as a monitoring tool during the post-diagnosis phase, we conducted association testing with clinical and pathological biomarkers in the PPMI and ADNI cohorts. We found a strong association between the imaging scores and cognitive measures, especially for the AD imaging score, and a weaker association for the PD imaging score. In addition, we observed a strong association between the imaging scores and the MoCA (P<0.001), MMSE (P<0.001), and ADAS-Cog-13 (P<0.001) scores, while the correlation with the MDS-UPDRS-III score was not as strong for the PD imaging score (**Figure 4C**). The imaging phenotypes can further help us assess the genetic architecture of neurodegenerative disorders. We found a significant association between imaging scores and disease-specific polygenic risk scores obtained from case-control genome-wide association studies (GWAS) in the UK Biobank cohort (**Figure 4D**). Finally, the screenshots of our cloud-based MRI prediction tool are shown in **Supplementary Figure 6 - 12**.

## Discussion

This study combines brain imaging data with machine learning to generate disease-specific scores. We have shown the potential of imaging scores in pre-diagnostic prediction, addressing heterogeneity and monitoring disease severity. Through model interpretation tools, we noticed that the hippocampus and temporal brain regions contribute the most to the ADRD score, while the substantia nigra contributes significantly to the PD score, reinforcing trust in our machine learning models. We observed that not all PD or all cause dementia subjects show abnormality in brain anatomical features during their pre-diagnostic phases. Finally, the association with clinical and pathological biomarkers shows that these scores can also be used to monitor disease progression. This study presents a general framework to generate machine learning based disease scores from complex data modalities such as imaging and is easily extendable to other disorders.

Detecting individuals in the asymptomatic phase is crucial for effective intervention with disease-modifying treatments. Given that brain atrophy initiates well before the onset of symptoms, utilizing structured brain imaging becomes essential. Our observation of significant variability in imaging suggests potential differences in underlying pathologies or co-pathology. Consequently, we posit that leveraging these scores could enhance intervention outcomes by ensuring a more homogeneous patient population at the commencement of a trial. In conjunction with pathological markers like α-synuclein seed amplification (Siderowf et al. 2023), imaging scores can serve as complementary measures, aiding in the precise selection of patients for inclusion in precision medicine trials. Similarly, these imaging scores can be used to boost the performance of multi-omics machine learning models (Makarious et al. 2022) for the early detection of ADRD and PD. Since the AUC of PD detection is lower compared to AD detection, we conclude that PD does not have a significant structured effect on the brain as captured by MRI. However, the incorporation of other risk factors such as age, sex, and polygenic risk scores enhances the performance of Parkinson’s disease prediction during the asymptomatic phase in UKB (see Table 3). Similar findings, depicting the importance of multimodal data for PD prediction, are shown by Makarious et al. in 2022. It’s important to note that there may be structural changes capable of distinguishing Parkinson’s Disease (PD) from healthy individuals, although higher resolution imaging is required.

Our models can be used as potential therapeutic engagement readouts. However, it is important to note that a single measure or modality may not be sufficient to capture all the symptoms of such disorders. Therefore, using multiple modalities (such as combining imaging, behavioral, and clinical data) could provide a more comprehensive and useful approach to assessing therapeutic engagement. Passive surveillance is another crucial aspect that has to be incorporated into health care systems for high-risk populations. It involves monitoring an individual’s data over time to objectively detect progression in risk even in the absence of symptoms. Given the increased availability of MRI in different clinical settings and its decreasing cost, our models have the potential to track patients’ brain health more effectively. Cognitive testing, though cheaper, is subjective and may not capture the full complexity of NDDs. Previous studies have indicated that NDDs encompass combinations of sleep, cognitive, and anxiety-related symptomatologies. MRI offers detailed structural and functional insights, providing objective measures for assessment. Alongside genomic data, MRI enhances diagnostic and predictive capabilities for NDDs. Our findings suggest that this integration offers a more comprehensive assessment compared to cognitive testing alone. Moreover, MRI allows for passive surveillance and transcends communication barriers, making it a versatile tool in NDD evaluation. Ultimately, this could result in improved patient triage, enhance the power of clinical trials, and lead to more efficient use of healthcare resources.

ML-based probabilities, like those proposed here, have proven useful in identifying new genetic loci for glaucoma and chronic obstructive pulmonary disease (Cosentino et al. 2023; Alipanahi et al. 2021). In our current analysis, we also showed an association of disease imaging scores with PRS of related disorders. This could have direct implications for imaging GWAS biobank-scale data. First, this may provide guidance toward understanding genetic risks that span neurodegenerative disorders. Second, these results may suggest endophenotypes that impact the brain and, ultimately, symptom onset. Third, the compressed representation provided by imaging scores may enable a more efficient search for novel risk loci, reducing proxy cases which is common in current GWAS studies.

Transparency and reproducibility is a critical aspect of science. It becomes furthermore important for machine learning models due to the dependency on data and the black box nature of machine learning models. To facilitate this, we have developed an interactive website that allows researchers to explore the top features contributing to predicting disease probabilities. Further, we incorporated a model perturbation analysis feature in our website where researchers can manually manipulate features and observe the resulting changes in imaging scores. Therefore, the transparency of our approach and the contributions of interpretable modeling move the research community away from black-box predictors.

Although this study presents the potential of machine learning based biomarkers on complex data such as imaging, much remains to be established. First, the limited availability of data from non-European participants’ datasets can introduce inherent bias. Second, the diagnosis that we have used both for validation and model training is not pathologically confirmed. We put efforts to avoid overfitting in our models so that the ML model won’t fit misdiagnosed cases. However, pathology-confirmed cases are particularly important for validation purposes. Third, in this work, we are considering ADRD as a single group but it is very important to have biomarkers that can differentiate between dementia types. Fourth, the analysis of common brain regions affected in both Alzheimer’s and Parkinson’s disease would be useful for future work. Finally, co-pathology is very common across neurodegenerative disorders which should be considered in future studies.

## Conclusions

In summary, this work presents an approach to generate disease biomarkers using machine learning and brain imaging data. Neurodegenerative and aging-related disorders are multi-system and remarkably complex. Our main conclusion, from both this work and our previous research, is that no single modality can fully capture the manifestations of these disorders. Models that integrate multiple modalities are showing the highest success. Thus, while imaging scores alone may bring limited added value, they are an important part of a multi-modal approach. This study is a step forward toward utilizing sophisticated machine-learning paradigms to facilitate the early disease detection, prognosis, and monitoring of disease progression.

To our knowledge, this is the first incidence of applying deep data from disease cohorts in imaging from time series data to surveillance-level studies at a biobank scale. We have also deployed this technology in an open science framework allowing users to submit images to a cloud-based platform that will empower them to explore and interpret their own data at little to no cost in under thirty minutes of processing time. The development of the first-of-its-kind cloud-based MRI prediction web application should lay the foundation for the upcoming generation of open-source digital tools in the biomedical community. Finally, the integration of these multimodal models for early detection can benefit trial recruitment, drug development, and clinical care practice.

## Data Availability

We have developed an interactive website (https://ndds-brainimaging-ml.streamlit.app) where researchers can investigate components of the predictive model and investigate feature effects on a sample and cohort level. To facilitate replication and expansion of our work, we have made the notebook publicly available on GitHub at https://github.com/NIH-CARD/NDDsImagingStreamlitApp and https://github.com/NIH-CARD/NDDsImaging. It includes all codes, figures, models, and supplements for this study. The code is part of the supplemental information; it includes the rendered Jupyter notebook with full step-by-step data preprocessing, statistical, and machine learning analyses.

## Acknowledgments

We thank the patients and their families who contributed to this research. This research was supported in part by the Intramural Research Program of the National Institute on Aging (NIA) and National Institute of Neurological Disorders and Stroke (NINDS), both part of the National Institutes of Health, within the Department of Health and Human Services; project number ZIA AG000534, ZO1 AG000949, ZIA-NS003154, and the Michael J Fox Foundation. Data used in the preparation of this article were obtained from the Parkinson’s Progression Markers Initiative (PPMI) database (www.ppmi-info.org/data). For up-to-date information on the study, visit www.ppmi-info.org. PPMI – a public-private partnership – is funded by the Michael J. Fox Foundation for Parkinson’s Research and funding partners, including Abbvie, Avid Radiopharmaceuticals, Biogen Idec, Bristol-Myers Squibb, Covance, Eli Lilly & Co., F. Hoffman-La Roche, Ltd., GE Healthcare, Genentech, GlaxoSmithKline, Lundbeck, Merck, MesoScale, Piramal, Pfizer, and UCB. Data and biospecimens used in the preparation of this manuscript were obtained from the Parkinson’s Disease Biomarkers Program (PDBP) Consortium, part of the National Institute of Neurological Disorders and Stroke at the National Institutes of Health. Investigators include: Roger Albin, Roy Alcalay, Alberto Ascherio, DuBois Bowman, Alice Chen-Plotkin, Ted Dawson, Richard Dewey, Dwight German, Xuemei Huang, Rachel Saunders-Pullman, Liana Rosenthal, Clemens Scherzer, David Vaillancourt, Vladislav Petyuk, Andy West and Jing Zhang. The PDBP Investigators have not participated in reviewing the data analysis or content of the manuscript.

Data collection and sharing for this project was funded by the Alzheimer’s Disease Neuroimaging Initiative (ADNI) (National Institutes of Health Grant U01 AG024904) and DOD ADNI (Department of Defense award number W81XWH-12-2-0012). ADNI is funded by the National Institute on Aging, the National Institute of Biomedical Imaging and Bioengineering, and through generous contributions from the following: AbbVie, Alzheimer’s Association; Alzheimer’s Drug Discovery Foundation; Araclon Biotech; BioClinica, Inc.; Biogen; Bristol-Myers Squibb Company; CereSpir, Inc.; Cogstate; Eisai Inc.; Elan Pharmaceuticals, Inc.; Eli Lilly and Company; EuroImmun; F. Hoffmann-La Roche Ltd and its affiliated company Genentech, Inc.; Fujirebio; GE Healthcare; IXICO Ltd.; Janssen Alzheimer Immunotherapy Research & Development, LLC.; Johnson & Johnson Pharmaceutical Research & Development LLC.; Lumosity; Lundbeck; Merck & Co., Inc.; Meso Scale Diagnostics, LLC.; NeuroRx Research; Neurotrack Technologies; Novartis Pharmaceuticals Corporation; Pfizer Inc.; Piramal Imaging; Servier; Takeda Pharmaceutical Company; and Transition Therapeutics. The Canadian Institutes of Health Research is providing funds to support ADNI clinical sites in Canada. Private sector contributions are facilitated by the Foundation for the National Institutes of Health (www.fnih.org). The grantee organization is the Northern California Institute for Research and Education, and the study is coordinated by the Alzheimer’s Therapeutic Research Institute at the University of Southern California.

## Author contributions

A.D., K.M., A.B.S., R.H.C., M.A.N., B.A. and F.F. contributed to the concept and design of the study. A.D., N.J.T., B.A. and F.F. were involved in the acquisition of data, data generation, and data cleaning. A.D., M.T., M.A.N., H.I., B.A. and F.F. did the analysis and interpretation of data. A.D., N.J.T., Ali D., A.B.S., M.A.N., H.I., B.A. and F.F. contributed to the drafting of the article and revising it critically.

