## supplement for "Prediction, prognosis and monitoring of neurodegeneration at biobank-scale via machine learning and imaging"

### Table of Contents

**Supplementary Material** **2**
Evaluation metrics 2
Feature importance and website development 2
Association testing 2
**Supplementary Figures** **4**
Supplementary Figure 1. Baseline age distribution of different cohorts grouped by the diagnosis status 4
Supplementary Figure 2. Procedure to train ensemble machine learning classifier using 3-fold cross-validation stratified based on individuals. 4
Supplementary Figure 3. Distribution of raw and transformed disease probabilities (using logit transformation, log (p / (1-p)) on different cohorts. 5
Supplementary Figure 4. Time to event distribution of subjects who converted to Dementia or PD after their image collection time point. We censored events for individuals with an attained survival of greater than 5 years (colored in orange). 5
Supplementary Figure 5. Diagnosis Age distribution for Dementia and PD patients in the UK biobank cohort. 6
Supplementary Figure 6. Home page for web application. 6
Supplementary Figure 7. Top discriminating brain MRI features of ADRD and PD using SHAP values. 7
Supplementary Figure 8. Users can upload a Nifti or DICOM file of their MRI image for analysis. 8
Supplementary Figure 9. Predicted probabilities of ADRD and PD for the MRI image uploaded by the user. 9
Supplementary Figure 10. The importance of different brain MRI regions in predicting probabilities of ADRD/PD. 10
Supplementary Figure 11. Force plot and decision plot illustrating the influence of each feature on the model's prediction for a single image uploaded by the user. 11
Supplementary Figure 12. Predicted probability and decision plot based on perturbed values using the What-If tool. 12
**Supplementary Tables** **13**
Supplementary Table 1. List of all feature extracted from brain imaging 13
Supplementary Table 2. Top hyperparameters for trained models and ensemble weights. 35

### Supplementary Material

#### Evaluation metrics

To evaluate the performance of Cox proportional hazards survival model, we used the following metrics:

- **Time-dependent AUC:** The receiver operating characteristic (ROC) curve and area under the curve (AUC) can be applied to survival data by defining sensitivity (true positive rate) and specificity (true negative rate) as time-dependent measures. In this context, cumulative cases refer to individuals who have experienced an event before or at a specified time, while dynamic controls are those who have not experienced the event by that time. The time-dependent AUC is used to determine how accurately a model can differentiate individuals who experience an event by a specific time point from those who experience it after that time. We used *sksurv.metrics.cumulative_dynamic_auc* function described here https://scikit-survival.readthedocs.io/.
- **C-index:** Concordance in this context refers to the correct ordering of two samples by the model, where the sample with a higher estimated risk score has a shorter actual survival time. If two samples have identical predicted risks, they are counted as concordant pairs with a weight of 0.5 instead of 1. We use the *sksurv.metrics.concordance_index_censored* function described here https://scikit-survival.readthedocs.io/.

#### Feature importance and website development

SHAP is an unified approach to explain the output of any supervised machine learning model. It assigns an importance value to every feature based on Shapley values. In addition, it generates the impact of each feature on the model's output i.e. the class probability for classification algorithms. We trained a surrogate LightGBM regression model (https://lightgbm.readthedocs.io/) using imaging features as input and disease probability scores (obtained from ensemble classifier) as output. We trained the surrogate model because of its compatibility with the SHAP package (https://shap.readthedocs.io/). To evaluate the contributing features, we use samples that are not involved in the ensemble model training. Imaging scores from surrogate models fits accurately with the scores obtained from original ensemble models (ADRD score: R-squared=0.87, PD score: R-squared=0.81). The SHAP package was used to calculate and visualize these Shapley values seen in the figures in the manuscript and the interactive website (https://ndds-brainimaging-ml.streamlit.app). To allow users to interrogate the model and evaluate its robustness we developed a what-if analysis tool using a reduced model that only uses top-20 features (available under the "Predict PD/ADRD disease" section of the website). We used force plot and decision plot to visualize individual predictions; for overall feature importance, beeswarm plot and bar plot were used (available in SHAP package). Finally, users can observe the interaction effects of different features for disease probability predictions using dependence plots.

#### Association testing

We utilized baseline data to test the association of imaging scores with clinical assessments and polygenic risk scores. For this purpose, we employed a linear regression model utilizing the statsmodels Python library. We adjusted for relevant covariates in our analysis.

- **with clinical and pathological biomarkers:** For clinical and biomarker assessment we use baseline data from ADNI and PPMI cohort. Also, we perform the analysis separately for each cohort (i.e., disease or control group) to avoid any possibility of confounder. We adjusted for age and sex in regression models. The formula is "Clinical Outcome ~ Imaging score + baseline_age + C(gender)". For meta analysis of all cohorts, we use the metafor package in R.
- **with polygenic risk scores:** We used UK biobank data to test for association between polygenic risk scores and imaging scores. We adjusted for age, sex, Townsend deprivation index and three genetic principal components available in the UK biobank database. The formula is "Imaging score ~ PRS + Townsend +

C(gender) + baseline_age + PC1 + PC2 + PC3". For meta analysis of all cohorts, we use the metafor package in R.

### Supplementary Figures

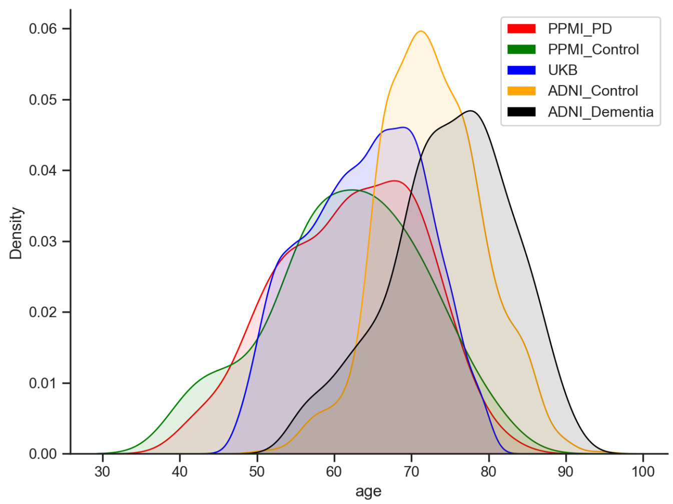


**Supplementary Figure 1. Baseline age distribution of different cohorts grouped by the diagnosis status**

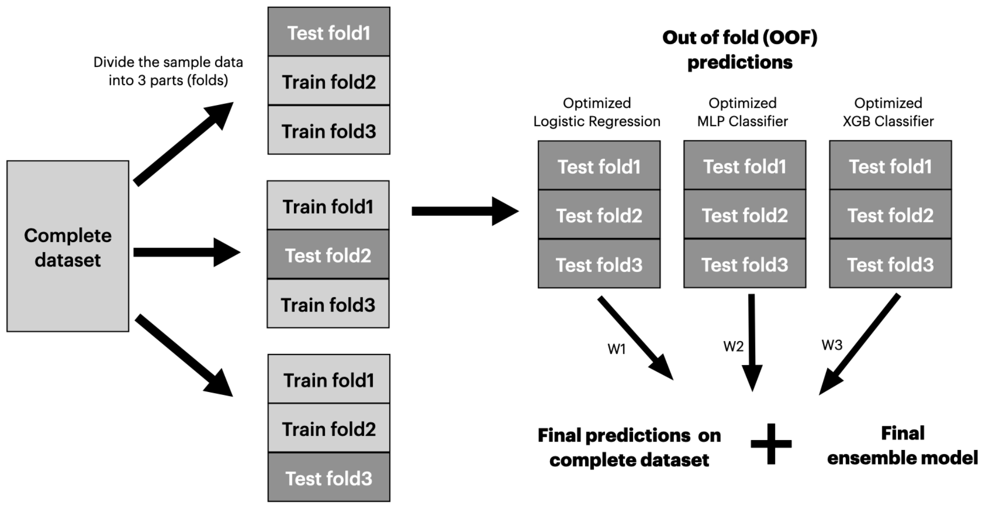


**Supplementary Figure 2. Procedure to train ensemble machine learning classifier using 3-fold cross-validation stratified based on individuals.**

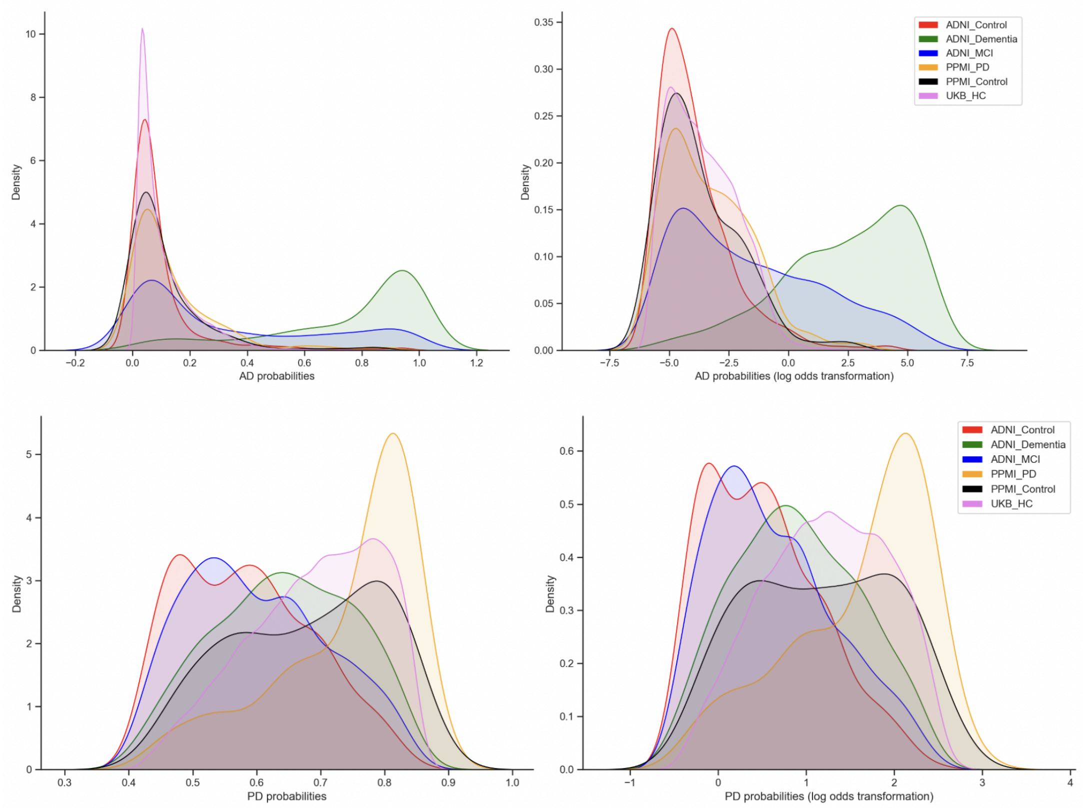


**Supplementary Figure 3. Distribution of raw and transformed disease probabilities (using logit transformation, *log (p / (1-p)*) on different cohorts.**

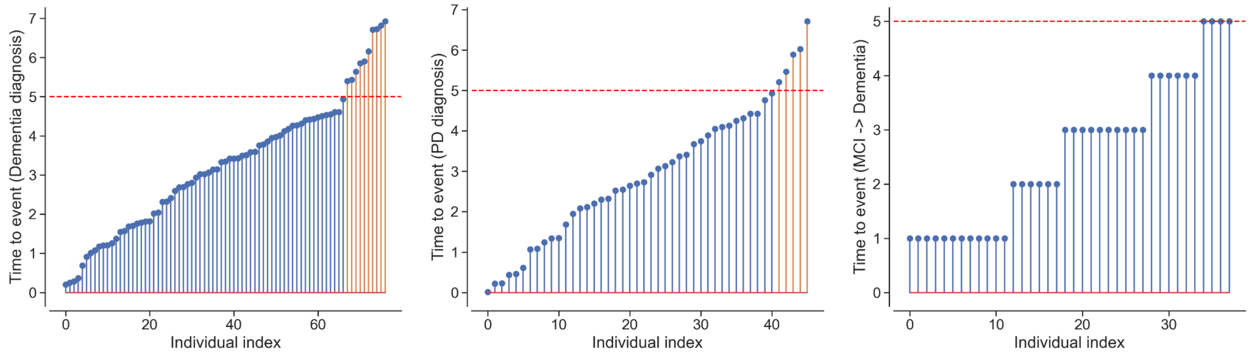


**Supplementary Figure 4. Time to event distribution of subjects who converted to Dementia or PD after their image collection time point. We censored events for individuals with an attained survival of greater than 5 years (colored in orange).**

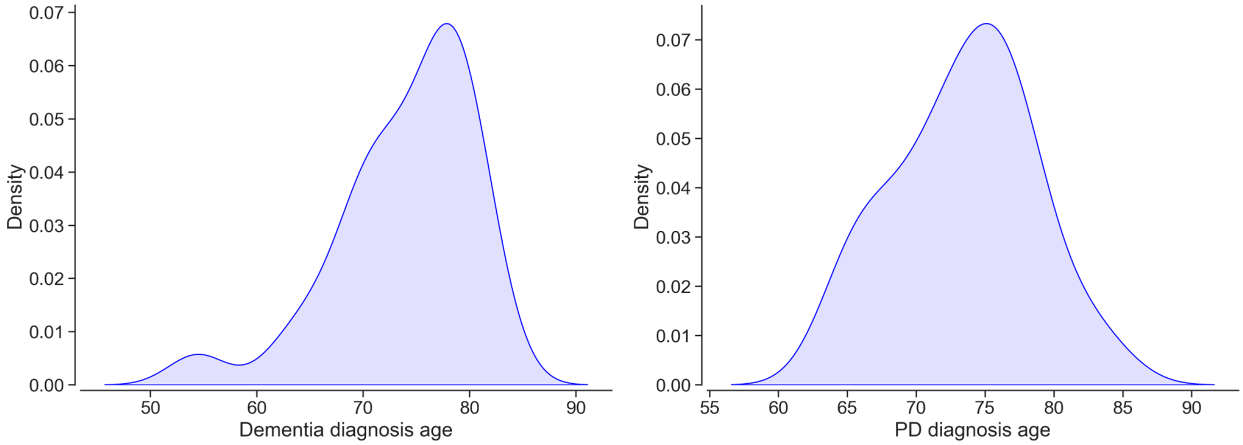


**Supplementary Figure 5. Diagnosis Age distribution for Dementia and PD patients in the UK biobank cohort.**

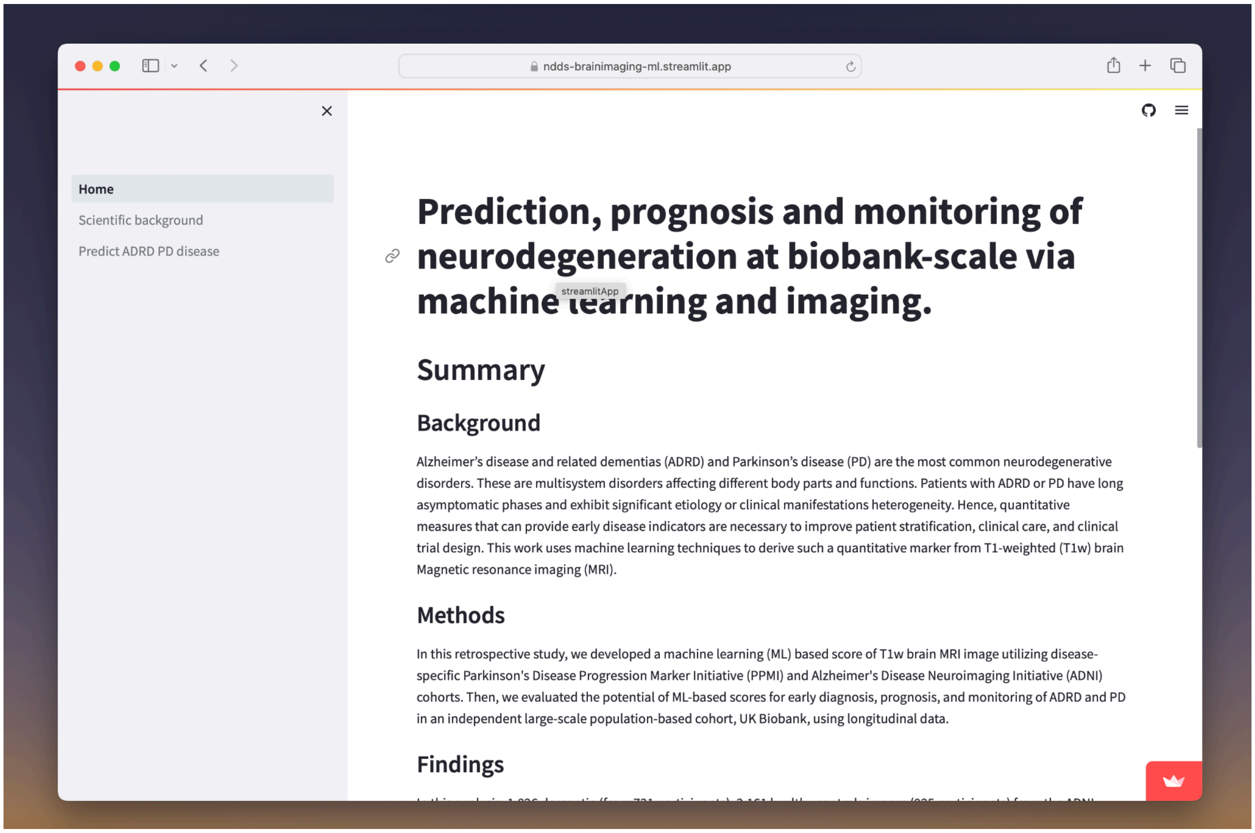


**Supplementary Figure 6. Home page for web application.**

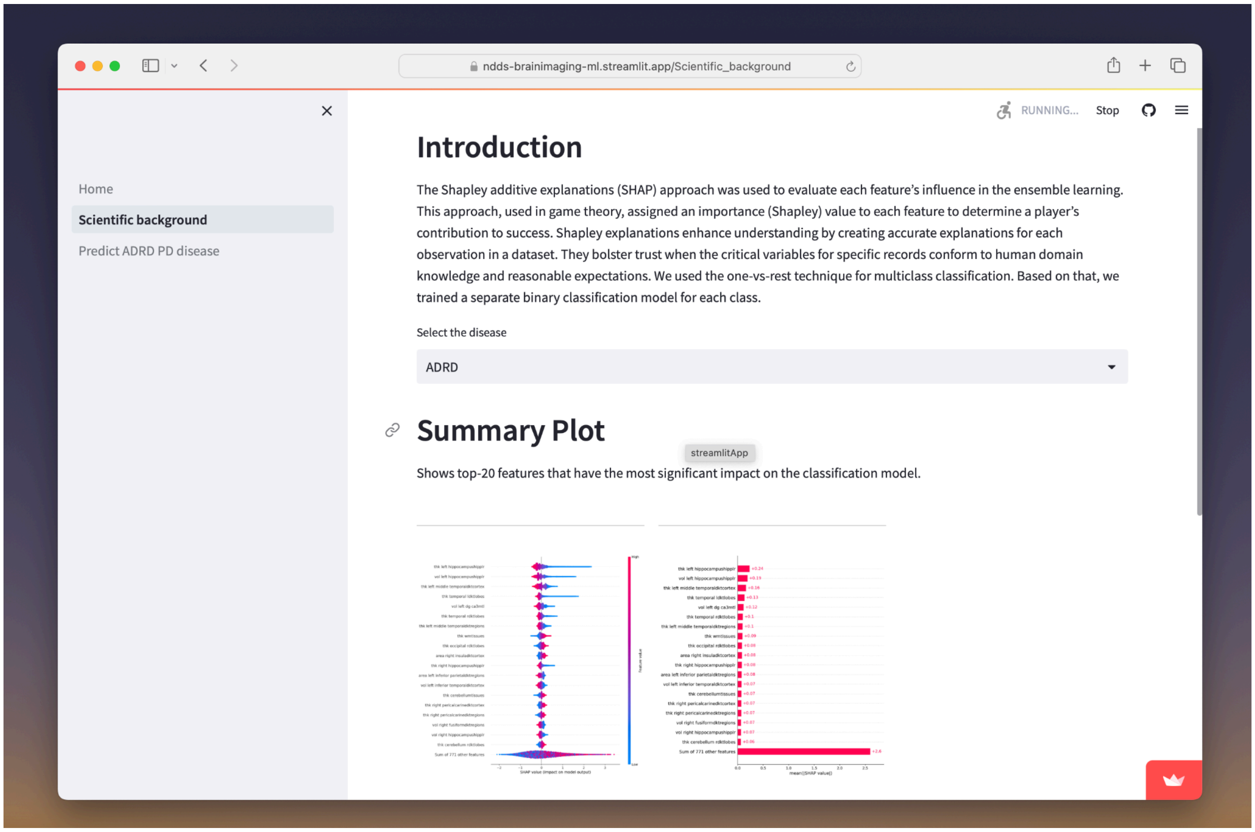


**Supplementary Figure 7. Top discriminating brain MRI features of ADRD and PD using SHAP values.**

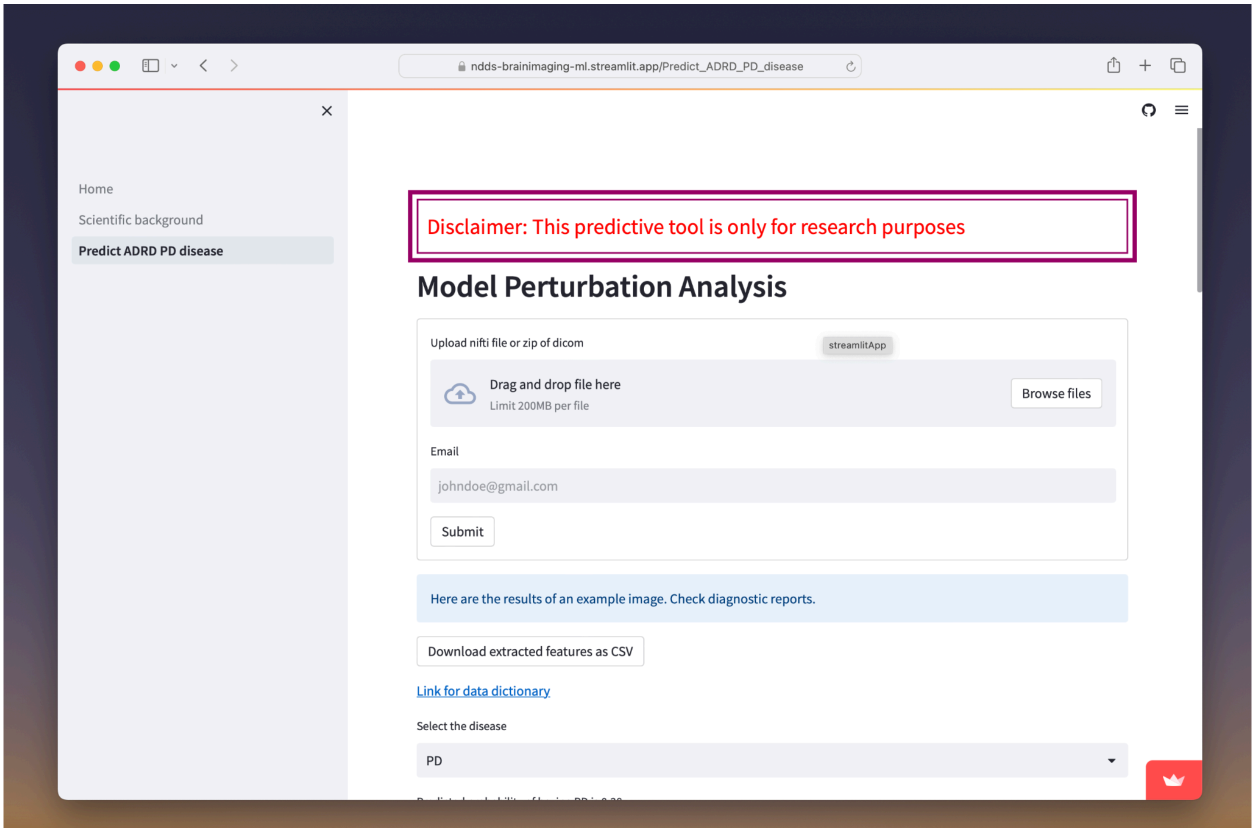


**Supplementary Figure 8. Users can upload a Nifti or DICOM file of their MRI image for analysis.**

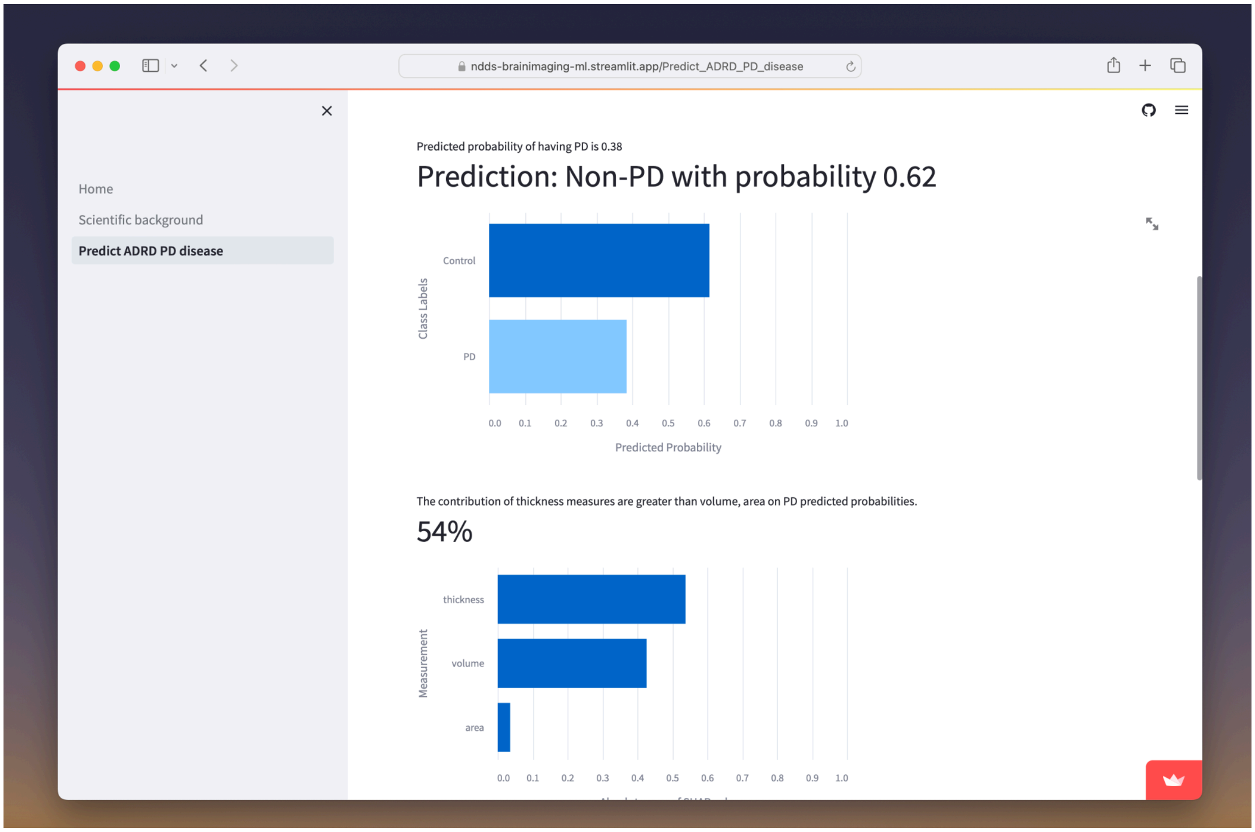


**Supplementary Figure 9. Predicted probabilities of ADRD and PD for the MRI image uploaded by the user.**

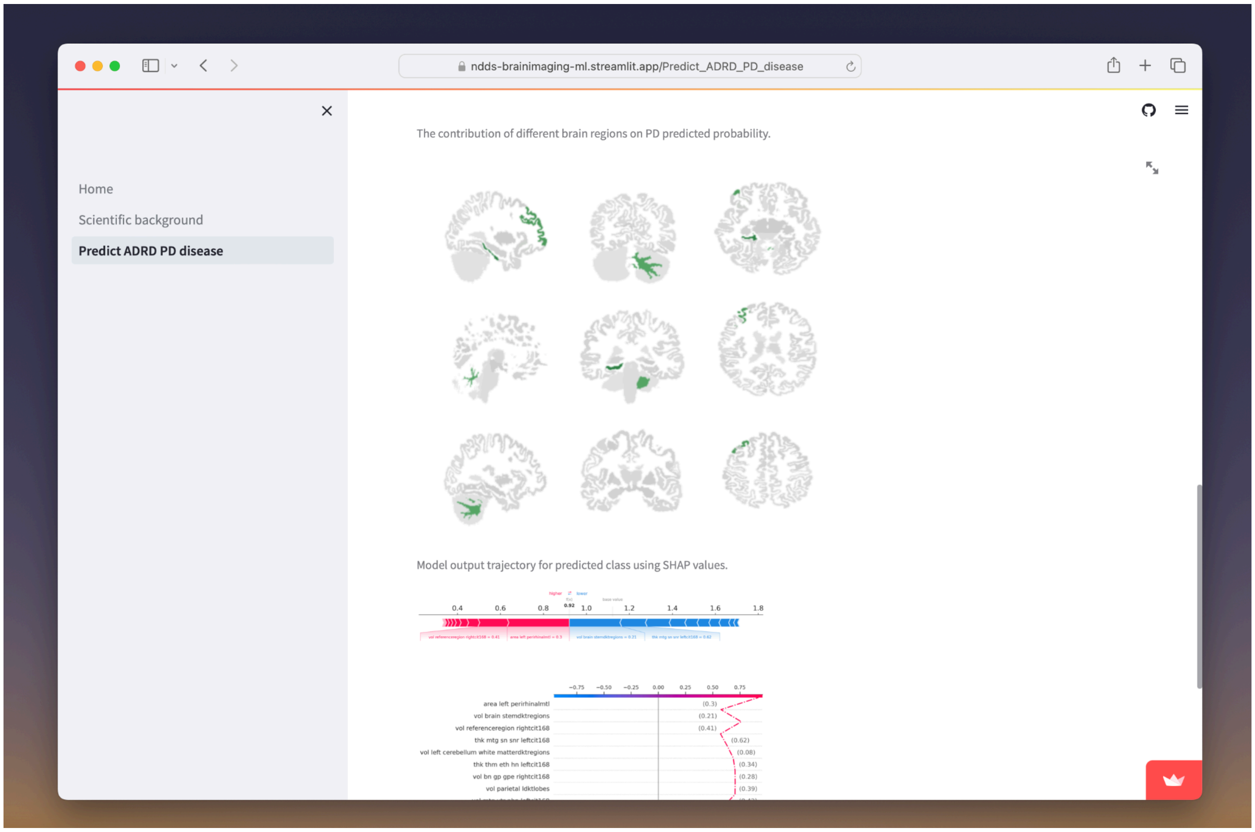


**Supplementary Figure 10. The importance of different brain MRI regions in predicting probabilities of ADRD/PD.**

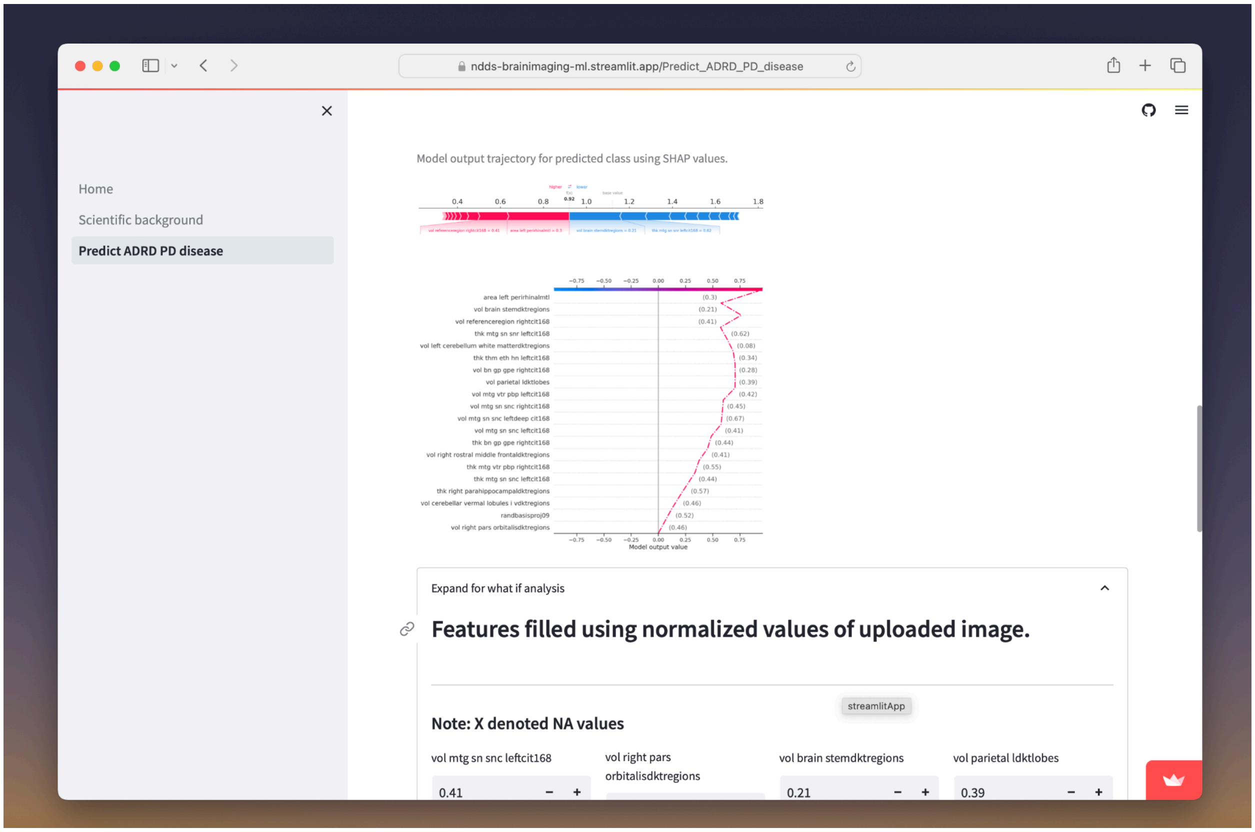


**Supplementary Figure 11. Force plot and decision plot illustrating the influence of each feature on the model's prediction for a single image uploaded by the user.**

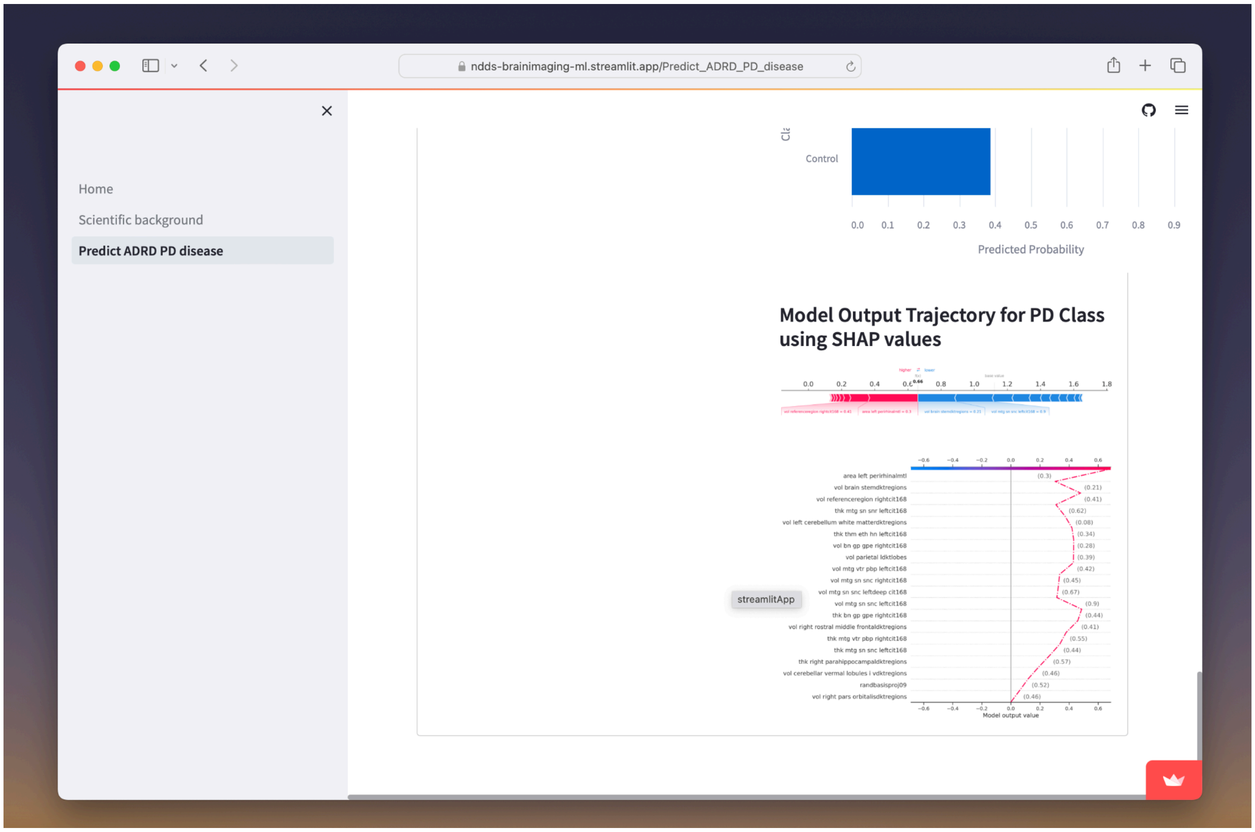


**Supplementary Figure 12. Predicted probability and decision plot based on perturbed values using the What-If tool.**

### Supplementary Tables

#### Supplementary Table 1. List of all feature extracted from brain imaging

| Feature ID | Missing data (%) | Included in ML model (Y/N) |
|---|---|---|
| thk_inferior_fronto_occipital_lwmtracts_right | 100 | N |
| thk_thm_eth_hn_leftdeep_cit168 | 100 | N |
| area_inferior_longitudinal_fasciculus_lwmtracts_right | 100 | N |
| area_mtg_vtr_vta_rightdeep_cit168 | 100 | N |
| thk_mtg_vtr_vta_rightdeep_cit168 | 100 | N |
| area_die_hth_mn_leftdeep_cit168 | 100 | N |
| thk_die_hth_mn_leftdeep_cit168 | 100 | N |
| thk_thm_eth_hn_rightdeep_cit168 | 100 | N |
| vol_mtg_vtr_vta_leftdeep_cit168 | 100 | N |
| area_thm_eth_hn_rightdeep_cit168 | 100 | N |
| thk_bn_gp_vep_rightdeep_cit168 | 100 | N |
| vol_die_sth_leftdeep_cit168 | 100 | N |
| area_mtg_vtr_pbp_rightdeep_cit168 | 100 | N |
| area_die_hth_leftdeep_cit168 | 100 | N |
| thk_die_sth_leftdeep_cit168 | 100 | N |
| vol_die_hth_mn_leftdeep_cit168 | 100 | N |
| area_die_sth_leftdeep_cit168 | 100 | N |
| thk_mtg_vtr_pbp_leftdeep_cit168 | 100 | N |
| vol_thm_eth_hn_leftdeep_cit168 | 100 | N |
| vol_mtg_vtr_vta_rightdeep_cit168 | 100 | N |
| thk_mtg_vtr_vta_leftdeep_cit168 | 100 | N |
| vol_uncinate_lwmtracts_right | 100 | N |
| area_inferior_fronto_occipital_lwmtracts_right | 100 | N |
| thk_bn_gp_vep_leftdeep_cit168 | 100 | N |
| vol_inferior_fronto_occipital_lwmtracts_right | 100 | N |
| area_uncinate_lwmtracts_right | 100 | N |
| thk_uncinate_lwmtracts_right | 100 | N |
| vol_die_hth_leftdeep_cit168 | 100 | N |
| area_bn_gp_vep_leftdeep_cit168 | 100 | N |
| area_mtg_vtr_vta_leftdeep_cit168 | 100 | N |
| thk_die_hth_leftdeep_cit168 | 100 | N |
| thk_mtg_vtr_pbp_rightdeep_cit168 | 100 | N |
| vol_bn_gp_vep_leftdeep_cit168 | 100 | N |
| thk_inferior_longitudinal_fasciculus_lwmtracts_right | 100 | N |

| | | |
|---|---|---|
| vol_mtg_vtr_pbp_rightdeep_cit168 | 100 | N |
| vol_inferior_longitudinal_fasciculus_lwmtracts_right | 100 | N |
| vol_die_sth_rightdeep_cit168 | 99.99 | N |
| area_die_sth_rightdeep_cit168 | 99.99 | N |
| area_die_hth_mn_rightdeep_cit168 | 99.99 | N |
| thk_die_hth_rightdeep_cit168 | 99.99 | N |
| area_die_hth_rightdeep_cit168 | 99.99 | N |
| vol_die_hth_rightdeep_cit168 | 99.99 | N |
| thk_die_sth_rightdeep_cit168 | 99.99 | N |
| vol_mtg_vtr_pbp_leftdeep_cit168 | 99.99 | N |
| vol_thm_eth_hn_rightdeep_cit168 | 99.99 | N |
| vol_die_hth_mn_rightdeep_cit168 | 99.99 | N |
| area_bn_gp_vep_rightdeep_cit168 | 99.99 | N |
| vol_bn_gp_vep_rightdeep_cit168 | 99.99 | N |
| area_mtg_vtr_pbp_leftdeep_cit168 | 99.99 | N |
| area_thm_eth_hn_leftdeep_cit168 | 99.99 | N |
| thk_die_hth_mn_rightdeep_cit168 | 99.99 | N |
| thk_corticospinal_lwmtracts_right | 99.89 | N |
| vol_corticospinal_lwmtracts_right | 99.88 | N |
| area_corticospinal_lwmtracts_right | 99.87 | N |
| vol_corticospinal_rwmtracts_left | 98.39 | N |
| thk_corticospinal_rwmtracts_left | 98.37 | N |
| area_corticospinal_rwmtracts_left | 98.36 | N |
| RandBasisProjPos05.1 | 96.92 | N |
| RandBasisProjPos03.1 | 96.92 | N |
| RandBasisProjPos08.1 | 96.92 | N |
| RandBasisProjPos10.1 | 96.92 | N |
| RandBasisProjPos07.1 | 96.92 | N |
| RandBasisProjPos09.1 | 96.92 | N |
| RandBasisProj09.1 | 96.92 | N |
| RandBasisProj08.1 | 96.92 | N |
| RandBasisProjPos04.1 | 96.92 | N |
| RandBasisProj07.1 | 96.92 | N |
| RandBasisProjPos06.1 | 96.92 | N |
| RandBasisProj05.1 | 96.92 | N |
| RandBasisProj02.1 | 96.92 | N |
| RandBasisProj01.1 | 96.92 | N |
| RandBasisProj03.1 | 96.92 | N |
| RandBasisProj10.1 | 96.92 | N |

| | | |
|---|---|---|
| RandBasisProj04.1 | 96.92 | N |
| RandBasisProjPos01.1 | 96.92 | N |
| RandBasisProjPos02.1 | 96.92 | N |
| RandBasisProj06.1 | 96.92 | N |
| thk_nbm_right_antbf | 17.39 | N |
| thk_nbm_right_midbf | 17.38 | N |
| thk_right_dg.ca3mtl | 0.47 | Y |
| vol_bn_gp_gpi_rightdeep_cit168 | 0.46 | Y |
| area_left_dg.ca3mtl | 0.46 | Y |
| thk_right_caudal_anterior_cingulatedktregions | 0.46 | Y |
| vol_right_dg.ca3mtl | 0.46 | Y |
| thk_right_pmecmtl | 0.46 | Y |
| vol_mtg_vtr_pbp_leftcit168 | 0.45 | Y |
| thk_left_dg.ca3mtl | 0.45 | Y |
| area_right_lateral_orbitofrontaldktregions | 0.45 | Y |
| vol_left_dg.ca3mtl | 0.45 | Y |
| area_ch13_leftbf | 0.45 | Y |
| area_inferior_fronto_occipital_lwmtracts_left | 0.45 | Y |
| vol_right_insuladktregions | 0.45 | Y |
| thk_corpus_callosumwmtracts_left | 0.45 | Y |
| area_right_precuneusdktregions | 0.45 | Y |
| vol_ch13_leftbf | 0.45 | Y |
| area_exa_rightcit168 | 0.45 | Y |
| thk_gmtissues | 0.45 | Y |
| area_right_dg.ca3mtl | 0.45 | Y |
| vol_right_pmecmtl | 0.45 | Y |
| area_bn_str_nac_rightcit168 | 0.45 | Y |
| thk_left_pmecmtl | 0.45 | Y |
| thk_mtg_sn_snr_leftdeep_cit168 | 0.45 | Y |
| vol_bn_str_nac_leftcit168 | 0.44 | Y |
| area_left_supramarginaldktregions | 0.44 | Y |
| thk_ch13_leftbf | 0.44 | Y |
| vol_referenceregion_rightcit168 | 0.44 | Y |
| thk_bn_str_pu_leftdeep_cit168 | 0.44 | Y |
| thk_mtg_sn_snr_leftcit168 | 0.44 | Y |
| thk_left_parahippocampalmtl | 0.44 | Y |
| area_right_lateral_occipitaldktregions | 0.44 | Y |
| area_right_precentraldktregions | 0.44 | Y |
| area_right_inferior_temporaldktregions | 0.44 | Y |

| | | |
|---|---|---|
| area_right_pars_opercularisdktregions | 0.44 | Y |
| thk_left_caudal_anterior_cingulatedktregions | 0.44 | Y |
| vol_referenceregion_rightdeep_cit168 | 0.44 | Y |
| vol_mtg_sn_snr_leftdeep_cit168 | 0.44 | Y |
| thk_parietal_rdktlobes | 0.44 | Y |
| vol_right_precuneusdktcortex | 0.44 | Y |
| vol_ch13_rightbf | 0.44 | Y |
| vol_right_pericalcarinedktregions | 0.44 | Y |
| vol_bn_str_ca_rightdeep_cit168 | 0.44 | Y |
| thk_brainstem_ldktlobes | 0.44 | Y |
| area_bn_gp_gpe_rightcit168 | 0.44 | Y |
| thk_left_lingualdktregions | 0.44 | Y |
| thk_bn_str_ca_leftdeep_cit168 | 0.44 | Y |
| vol_right_lateral_orbitofrontaldktregions | 0.44 | Y |
| vol_right_cuneusdktcortex | 0.44 | Y |
| area_nbm_right_midbf | 0.44 | Y |
| thk_bn_str_ca_rightcit168 | 0.44 | Y |
| vol_right_postcentraldktcortex | 0.44 | Y |
| area_die_sth_rightcit168 | 0.44 | Y |
| thk_left_inferior_temporaldktregions | 0.44 | Y |
| vol_left_rostral_middle_frontaldktregions | 0.44 | Y |
| vol_right_lingualdktregions | 0.44 | Y |
| area_right_subiculummtl | 0.44 | Y |
| area_right_lateral_occipitaldktcortex | 0.44 | Y |
| area_bn_str_pu_rightcit168 | 0.44 | Y |
| thk_mtg_rn_leftcit168 | 0.44 | Y |
| area_right_lingualdktcortex | 0.44 | Y |
| wmh_vol | 0.44 | Y |
| area_left_middle_temporaldktregions | 0.44 | Y |
| vol_cerebellumtissues | 0.44 | Y |
| vol_die_hth_mn_leftcit168 | 0.44 | Y |
| thk_uncinate_lwmtracts_left | 0.44 | Y |
| area_bn_str_pu_leftcit168 | 0.44 | Y |
| thk_uncinate_rwmtracts_right | 0.44 | Y |
| area_right_pars_opercularisdktcortex | 0.44 | Y |
| thk_right_superior_frontaldktregions | 0.44 | Y |
| vol_die_hth_leftcit168 | 0.44 | Y |
| thk_right_transverse_temporaldktcortex | 0.44 | Y |
| thk_mtg_sn_snc_leftcit168 | 0.44 | Y |

| | | |
|---|---|---|
| thk_left_paracentraldktcortex | 0.44 | Y |
| vol_right_medial_orbitofrontaldktregions | 0.44 | Y |
| vol_left_supramarginaldktregions | 0.44 | Y |
| vol_right_alecmtl | 0.44 | Y |
| thk_nbm_left_posbf | 0.44 | Y |
| vol_left_inferior_temporaldktcortex | 0.44 | Y |
| area_die_hth_mn_leftcit168 | 0.44 | Y |
| vol_right_postcentraldktregions | 0.44 | Y |
| thk_right_supramarginaldktcortex | 0.44 | Y |
| thk_nbm_left_antbf | 0.44 | Y |
| thk_bn_gp_gpe_leftcit168 | 0.44 | Y |
| vol_bn_gp_gpe_leftdeep_cit168 | 0.44 | Y |
| thk_brainstem_rdktlobes | 0.44 | Y |
| thk_left_caudal_middle_frontaldktcortex | 0.44 | Y |
| area_left_pmecmtl | 0.44 | Y |
| area_bn_gp_vep_leftcit168 | 0.44 | Y |
| thk_occipital_ldktlobes | 0.44 | Y |
| area_right_superior_parietaldktcortex | 0.44 | Y |
| vol_bn_gp_gpe_rightcit168 | 0.44 | Y |
| thk_right_superior_parietaldktcortex | 0.44 | Y |
| thk_left_superior_parietaldktregions | 0.44 | Y |
| vol_right_middle_temporaldktcortex | 0.44 | Y |
| area_left_pallidiumdktregions | 0.44 | Y |
| area_mtg_sn_snc_rightdeep_cit168 | 0.44 | Y |
| area_left_fusiformdktcortex | 0.44 | Y |
| area_left_posterior_cingulatedktcortex | 0.44 | Y |
| area_temporal_ldktlobes | 0.44 | Y |
| thk_right_subiculummtl | 0.44 | Y |
| vol_left_caudal_middle_frontaldktregions | 0.44 | Y |
| thk_cerebellum_ldktlobes | 0.44 | Y |
| vol_right_lateral_occipitaldktcortex | 0.44 | Y |
| area_right_pars_triangularisdktregions | 0.44 | Y |
| area_bn_gp_vep_rightcit168 | 0.44 | Y |
| area_bn_str_ca_leftdeep_cit168 | 0.44 | Y |
| vol_right_fusiformdktregions | 0.44 | Y |
| thk_right_amygdaladktregions | 0.44 | Y |
| thk_left_ventral_dcdktregions | 0.44 | Y |
| thk_mtg_rn_rightdeep_cit168 | 0.44 | Y |
| area_uncinate_lwmtracts_left | 0.44 | Y |

| | | |
|---|---|---|
| vol_right_putamendktregions | 0.44 | Y |
| area_left_superior_parietaldktregions | 0.44 | Y |
| thk_bn_gp_gpi_rightcit168 | 0.44 | Y |
| area_left_lingualdktregions | 0.44 | Y |
| thk_left_posterior_cingulatedktcortex | 0.44 | Y |
| area_superior_longitudinal_fasciculus_rwmtracts_right | 0.44 | Y |
| thk_right_cerebellum_exteriordktregions | 0.44 | Y |
| vol_die_hth_rightcit168 | 0.44 | Y |
| thk_right_perirhinalmtl | 0.44 | Y |
| thk_bn_gp_vep_rightcit168 | 0.44 | Y |
| area_exa_leftcit168 | 0.44 | Y |
| thk_bn_str_ca_leftcit168 | 0.44 | Y |
| vol_bn_str_nac_rightcit168 | 0.44 | Y |
| vol_bn_str_ca_leftcit168 | 0.44 | Y |
| area_left_superior_temporaldktcortex | 0.44 | Y |
| thk_left_insuladktregions | 0.44 | Y |
| area_inferior_longitudinal_fasciculus_rwmtracts_right | 0.43 | Y |
| area_deepgraytissues | 0.43 | Y |
| area_left_ca1mtl | 0.43 | Y |
| thk_left_fusiformdktregions | 0.43 | Y |
| thk_nbm_right_posbf | 0.43 | Y |
| vol_right_isthmus_cingulatedktregions | 0.43 | Y |
| vol_bn_gp_gpe_leftcit168 | 0.43 | Y |
| vol_mtg_sn_snc_rightcit168 | 0.43 | Y |
| thk_left_medial_orbitofrontaldktcortex | 0.43 | Y |
| area_right_posterior_cingulatedktcortex | 0.43 | Y |
| thk_right_rostral_middle_frontaldktcortex | 0.43 | Y |
| area_right_lateral_ventricledktregions | 0.43 | Y |
| area_right_rostral_middle_frontaldktregions | 0.43 | Y |
| area_left_parahippocampalmtl | 0.43 | Y |
| vol_wmtissues | 0.43 | Y |
| vol_right_parahippocampalmtl | 0.43 | Y |
| thk_left_alecmtl | 0.43 | Y |
| area_right_parahippocampalmtl | 0.43 | Y |
| area_left_pars_opercularisdktcortex | 0.43 | Y |
| thk_inferior_fronto_occipital_lwmtracts_left | 0.43 | Y |
| area_left_rostral_middle_frontaldktregions | 0.43 | Y |
| vol_nbm_left_midbf | 0.43 | Y |
| area_left_lingualdktcortex | 0.43 | Y |

| | | |
|---|---|---|
| area_bn_str_ca_rightdeep_cit168 | 0.43 | Y |
| area_left_thalamus_properdktregions | 0.43 | Y |
| area_left_pericalcarinedktcortex | 0.43 | Y |
| area_nbm_right_posbf | 0.43 | Y |
| vol_mtg_sn_snr_leftcit168 | 0.43 | Y |
| area_right_superior_frontaldktcortex | 0.43 | Y |
| area_left_cerebellum_white_matterdktregions | 0.43 | Y |
| vol_left_lingualdktcortex | 0.43 | Y |
| vol_left_inferior_parietaldktcortex | 0.43 | Y |
| vol_right_perirhinalmtl | 0.43 | Y |
| vol_die_sth_leftcit168 | 0.43 | Y |
| area_die_hth_rightcit168 | 0.43 | Y |
| vol_left_lateral_orbitofrontaldktregions | 0.43 | Y |
| area_mtg_sn_snr_rightdeep_cit168 | 0.43 | Y |
| vol_left_superior_temporaldktcortex | 0.43 | Y |
| thk_inferior_longitudinal_fasciculus_rwmtracts_right | 0.43 | Y |
| thk_right_rostral_anterior_cingulatedktcortex | 0.43 | Y |
| area_left_medial_orbitofrontaldktregions | 0.43 | Y |
| thk_cerebellumtissues | 0.43 | Y |
| area_left_subiculummtl | 0.43 | Y |
| thk_mtg_sn_snc_rightdeep_cit168 | 0.43 | Y |
| thk_corpus_callosumwmtracts_right | 0.43 | Y |
| area_right_fusiformdktcortex | 0.43 | Y |
| thk_bn_gp_gpe_leftdeep_cit168 | 0.43 | Y |
| thk_bn_gp_gpi_leftdeep_cit168 | 0.43 | Y |
| area_mtg_vtr_vta_leftcit168 | 0.43 | Y |
| area_left_amygdaladktregions | 0.43 | Y |
| area_mtg_vtr_vta_rightcit168 | 0.43 | Y |
| vol_right_precuneusdktregions | 0.43 | Y |
| vol_mtg_rn_rightdeep_cit168 | 0.43 | Y |
| thk_left_middle_temporaldktcortex | 0.43 | Y |
| vol_left_pars_opercularisdktcortex | 0.43 | Y |
| thk_right_ca1mtl | 0.43 | Y |
| area_left_entorhinaldktcortex | 0.43 | Y |
| area_inferior_longitudinal_fasciculus_lwmtracts_left | 0.43 | Y |
| thk_left_inferior_parietaldktcortex | 0.43 | Y |
| area_temporal_rdktlobes | 0.43 | Y |
| area_bn_str_pu_leftdeep_cit168 | 0.43 | Y |
| area_left_isthmus_cingulatedktregions | 0.43 | Y |

| | | |
|---|---|---|
| thk_right_cuneusdktcortex | 0.43 | Y |
| vol_brainstem_rdktlobes | 0.43 | Y |
| area_left_superior_frontaldktcortex | 0.43 | Y |
| vol_cerebellar_vermal_lobules_vi.viidktregions | 0.43 | Y |
| area_left_lateral_ventricledktregions | 0.43 | Y |
| thk_deepgraytissues | 0.43 | Y |
| vol_referenceregion_leftdeep_cit168 | 0.43 | Y |
| area_mtg_vtr_pbp_rightcit168 | 0.43 | Y |
| area_left_pericalcarinedktregions | 0.43 | Y |
| thk_left_transverse_temporaldktcortex | 0.43 | Y |
| thk_corticospinal_rwmtracts_right | 0.43 | Y |
| vol_right_insuladktcortex | 0.43 | Y |
| thk_right_putamendktregions | 0.43 | Y |
| thk_right_caudatedktregions | 0.43 | Y |
| area_right_hippocampushipplr | 0.43 | Y |
| thk_right_isthmus_cingulatedktregions | 0.43 | Y |
| area_bn_gp_gpi_rightdeep_cit168 | 0.43 | Y |
| vol_inferior_longitudinal_fasciculus_rwmtracts_right | 0.43 | Y |
| area_left_insuladktcortex | 0.43 | Y |
| vol_csfdktregions | 0.43 | Y |
| thk_mtg_vtr_pbp_rightcit168 | 0.43 | Y |
| thk_left_postcentraldktcortex | 0.43 | Y |
| area_brainstemtissues | 0.43 | Y |
| thk_right_postcentraldktcortex | 0.43 | Y |
| thk_right_middle_temporaldktregions | 0.43 | Y |
| vol_left_lateral_occipitaldktregions | 0.43 | Y |
| vol_right_caudal_anterior_cingulatedktcortex | 0.43 | Y |
| thk_corticospinal_lwmtracts_left | 0.43 | Y |
| thk_mtg_vtr_vta_leftcit168 | 0.43 | Y |
| area_left_rostral_anterior_cingulatedktregions | 0.43 | Y |
| vol_uncinate_lwmtracts_left | 0.43 | Y |
| area_referenceregion_rightdeep_cit168 | 0.43 | Y |
| area_left_perirhinalmtl | 0.43 | Y |
| thk_bn_str_ca_rightdeep_cit168 | 0.43 | Y |
| vol_right_pars_orbitalisdktcortex | 0.43 | Y |
| area_left_cuneusdktcortex | 0.43 | Y |
| area_left_hippocampushipplr | 0.43 | Y |
| vol_right_pars_triangularisdktregions | 0.43 | Y |
| thk_cerebellar_vermal_lobules_i.vdktregions | 0.43 | Y |

| | | |
|---|---|---|
| area_right_pars_triangularisdktcortex | 0.43 | Y |
| vol_right_supramarginaldktregions | 0.43 | Y |
| vol_left_fusiformdktcortex | 0.43 | Y |
| area_mtg_sn_snr_leftsnseg | 0.43 | Y |
| thk_left_inferior_parietaldktregions | 0.43 | Y |
| thk_right_inferior_parietaldktregions | 0.43 | Y |
| thk_mtg_sn_snr_rightcit168 | 0.43 | Y |
| vol_left_thalamus_properdktregions | 0.43 | Y |
| area_right_ventral_dcdktregions | 0.43 | Y |
| thk_left_precuneusdktcortex | 0.43 | Y |
| vol_exa_rightcit168 | 0.43 | Y |
| thk_right_lateral_orbitofrontaldktregions | 0.43 | Y |
| vol_right_ca1mtl | 0.43 | Y |
| vol_cerebellar_vermal_lobules_viii.xdktregions | 0.43 | Y |
| thk_cerebellar_vermal_lobules_viii.xdktregions | 0.43 | Y |
| area_left_pars_triangularisdktregions | 0.43 | Y |
| vol_left_precuneusdktcortex | 0.43 | Y |
| vol_superior_longitudinal_fasciculus_rwmtracts_right | 0.43 | Y |
| area_right_pericalcarinedktregions | 0.43 | Y |
| thk_bn_gp_vep_leftcit168 | 0.43 | Y |
| thk_right_precuneusdktcortex | 0.43 | Y |
| vol_nbm_left_posbf | 0.43 | Y |
| area_left_lateral_orbitofrontaldktcortex | 0.43 | Y |
| vol_left_precentraldktregions | 0.43 | Y |
| area_right_cerebellum_exteriordktregions | 0.43 | Y |
| vol_left_pars_opercularisdktregions | 0.43 | Y |
| thk_bn_str_nac_leftcit168 | 0.43 | Y |
| area_right_insuladktregions | 0.43 | Y |
| thk_wmtissues | 0.43 | Y |
| area_right_paracentraldktcortex | 0.43 | Y |
| area_right_middle_temporaldktregions | 0.43 | Y |
| area_ch13_rightbf | 0.43 | Y |
| area_corpus_callosumwmtracts_right | 0.43 | Y |
| vol_left_middle_temporaldktcortex | 0.43 | Y |
| vol_right_pars_opercularisdktregions | 0.43 | Y |
| vol_nbm_right_antbf | 0.43 | Y |
| thk_right_entorhinaldktcortex | 0.43 | Y |
| vol_right_pericalcarinedktcortex | 0.43 | Y |
| thk_left_superior_temporaldktregions | 0.43 | Y |

| | | |
|---|---|---|
| thk_right_pars_opercularisdktcortex | 0.43 | Y |
| thk_referenceregion_rightdeep_cit168 | 0.43 | Y |
| vol_right_rostral_anterior_cingulatedktregions | 0.43 | Y |
| area_left_putamendktregions | 0.43 | Y |
| area_bn_gp_gpi_leftcit168 | 0.43 | Y |
| vol_left_paracentraldktregions | 0.43 | Y |
| thk_left_lateral_orbitofrontaldktregions | 0.43 | Y |
| area_left_postcentraldktregions | 0.43 | Y |
| area_right_isthmus_cingulatedktcortex | 0.43 | Y |
| vol_left_alecmtl | 0.43 | Y |
| thk_right_superior_parietaldktregions | 0.43 | Y |
| vol_left_paracentraldktcortex | 0.43 | Y |
| vol_bn_gp_vep_leftcit168 | 0.43 | Y |
| vol_left_insuladktregions | 0.43 | Y |
| vol_right_entorhinaldktcortex | 0.43 | Y |
| thk_right_posterior_cingulatedktcortex | 0.43 | Y |
| vol_left_isthmus_cingulatedktcortex | 0.43 | Y |
| vol_brainstemtissues | 0.43 | Y |
| thk_right_precuneusdktregions | 0.43 | Y |
| vol_thm_eth_hn_rightcit168 | 0.43 | Y |
| area_left_cerebellem_exteriordktregions | 0.43 | Y |
| vol_left_pericalcarinedktregions | 0.43 | Y |
| thk_die_hth_mn_leftcit168 | 0.43 | Y |
| vol_csftissues | 0.43 | Y |
| vol_temporal_rdktlobes | 0.43 | Y |
| thk_left_parahippocampaldktcortex | 0.43 | Y |
| area_right_precentraldktcortex | 0.43 | Y |
| area_cerebellum_rdktlobes | 0.43 | Y |
| vol_bn_str_ca_leftdeep_cit168 | 0.43 | Y |
| area_die_hth_leftcit168 | 0.43 | Y |
| thk_left_putamendktregions | 0.43 | Y |
| thk_bn_str_pu_leftcit168 | 0.43 | Y |
| vol_right_transverse_temporaldktregions | 0.43 | Y |
| area_left_caudal_anterior_cingulatedktregions | 0.43 | Y |
| vol_right_pars_opercularisdktcortex | 0.43 | Y |
| area_nbm_left_posbf | 0.43 | Y |
| vol_4th_ventricledktregions | 0.43 | Y |
| vol_corticospinal_lwmtracts_left | 0.43 | Y |
| area_left_entorhinaldktregions | 0.43 | Y |

| | | |
|---|---|---|
| thk_left_pars_opercularisdktcortex | 0.43 | Y |
| area_4th_ventricledktregions | 0.43 | Y |
| area_die_hth_mn_rightcit168 | 0.43 | Y |
| area_corpus_callosumwmtracts_left | 0.43 | Y |
| area_left_pars_orbitalisdktregions | 0.43 | Y |
| area_csfdktregions | 0.43 | Y |
| thk_bn_gp_gpi_leftcit168 | 0.43 | Y |
| vol_occipital_rdktlobes | 0.43 | Y |
| thk_left_pars_triangularisdktregions | 0.43 | Y |
| thk_right_caudal_middle_frontaldktcortex | 0.43 | Y |
| thk_ch13_rightbf | 0.43 | Y |
| thk_left_middle_temporaldktregions | 0.43 | Y |
| vol_bn_gp_vep_rightcit168 | 0.43 | Y |
| area_nbm_left_antbf | 0.43 | Y |
| vol_left_pmecmtl | 0.43 | Y |
| vol_right_caudal_middle_frontaldktcortex | 0.43 | Y |
| thk_thm_eth_hn_leftcit168 | 0.43 | Y |
| thk_left_thalamus_properdktregions | 0.43 | Y |
| thk_mtg_vtr_pbp_leftcit168 | 0.43 | Y |
| area_right_pmecmtl | 0.43 | Y |
| thk_right_thalamus_properdktregions | 0.43 | Y |
| vol_bn_str_ca_rightcit168 | 0.43 | Y |
| thk_left_isthmus_cingulatedktcortex | 0.43 | Y |
| area_nbm_right_antbf | 0.43 | Y |
| area_right_caudal_anterior_cingulatedktcortex | 0.43 | Y |
| thk_right_superior_temporaldktregions | 0.43 | Y |
| area_bn_gp_gpi_leftdeep_cit168 | 0.43 | Y |
| thk_left_lateral_occipitaldktcortex | 0.43 | Y |
| vol_left_hippocampusdktregions | 0.43 | Y |
| thk_left_isthmus_cingulatedktregions | 0.43 | Y |
| vol_nbm_right_posbf | 0.43 | Y |
| vol_left_parahippocampaldktregions | 0.43 | Y |
| thk_left_rostral_anterior_cingulatedktcortex | 0.43 | Y |
| wmh_log_evidence | 0.43 | Y |
| vol_bn_str_pu_leftdeep_cit168 | 0.43 | Y |
| area_left_precentraldktcortex | 0.43 | Y |
| area_left_lateral_occipitaldktregions | 0.43 | Y |
| thk_right_cerebellum_white_matterdktregions | 0.43 | Y |
| vol_mtg_vtr_pbp_rightcit168 | 0.43 | Y |

| | | |
|---|---|---|
| area_right_supramarginaldktregions | 0.43 | Y |
| vol_right_parahippocampaldktcortex | 0.43 | Y |
| thk_left_fusiformdktcortex | 0.43 | Y |
| thk_right_lingualdktcortex | 0.43 | Y |
| thk_mtg_rn_rightcit168 | 0.43 | Y |
| area_bn_gp_gpe_rightdeep_cit168 | 0.43 | Y |
| area_right_caudal_middle_frontaldktcortex | 0.43 | Y |
| thk_right_parahippocampaldktregions | 0.43 | Y |
| vol_corticospinal_rwmtracts_right | 0.43 | Y |
| vol_bn_gp_gpi_rightcit168 | 0.43 | Y |
| thk_left_cuneusdktcortex | 0.43 | Y |
| area_left_precentraldktregions | 0.43 | Y |
| thk_right_pars_triangularisdktregions | 0.43 | Y |
| vol_exa_leftcit168 | 0.43 | Y |
| thk_cerebellar_vermal_lobules_vi.viidktregions | 0.43 | Y |
| vol_nbm_left_antbf | 0.43 | Y |
| vol_right_isthmus_cingulatedktcortex | 0.43 | Y |
| area_left_precuneusdktregions | 0.43 | Y |
| thk_frontal_ldktlobes | 0.43 | Y |
| vol_left_precuneusdktregions | 0.43 | Y |
| area_bn_gp_gpe_leftdeep_cit168 | 0.43 | Y |
| thk_right_pars_opercularisdktregions | 0.43 | Y |
| thk_csfdktregions | 0.43 | Y |
| thk_right_lingualdktregions | 0.43 | Y |
| thk_right_pericalcarinedktcortex | 0.43 | Y |
| thk_bn_gp_gpe_rightdeep_cit168 | 0.43 | Y |
| thk_left_caudatedktregions | 0.43 | Y |
| thk_left_pars_triangularisdktcortex | 0.43 | Y |
| vol_left_pars_triangularisdktregions | 0.43 | Y |
| area_right_pars_orbitalisdktregions | 0.43 | Y |
| vol_mtg_vtr_vta_leftcit168 | 0.43 | Y |
| thk_mtg_vtr_vta_rightcit168 | 0.43 | Y |
| area_left_caudal_anterior_cingulatedktcortex | 0.43 | Y |
| vol_right_pars_triangularisdktcortex | 0.43 | Y |
| vol_frontal_ldktlobes | 0.43 | Y |
| thk_right_precentraldktregions | 0.43 | Y |
| thk_right_alecmtl | 0.43 | Y |
| thk_referenceregion_leftdeep_cit168 | 0.43 | Y |
| area_cerebellar_vermal_lobules_i.vdktregions | 0.43 | Y |

| | | |
|---|---|---|
| thk_left_subiculummtl | 0.43 | Y |
| area_mtg_rn_rightdeep_cit168 | 0.42 | Y |
| thk_superior_longitudinal_fasciculus_lwmtracts_left | 0.42 | Y |
| thk_left_hippocampushipplr | 0.42 | Y |
| thk_left_medial_orbitofrontaldktregions | 0.42 | Y |
| thk_left_parahippocampaldktregions | 0.42 | Y |
| thk_right_pars_orbitalisdktregions | 0.42 | Y |
| area_left_inferior_temporaldktregions | 0.42 | Y |
| area_right_fusiformdktregions | 0.42 | Y |
| area_thm_eth_hn_rightcit168 | 0.42 | Y |
| area_right_postcentraldktcortex | 0.42 | Y |
| area_right_thalamus_properdktregions | 0.42 | Y |
| thk_left_precentraldktregions | 0.42 | Y |
| thk_left_cerebellum_white_matterdktregions | 0.42 | Y |
| vol_corpus_callosumwmtracts_left | 0.42 | Y |
| area_brain_stemdktregions | 0.42 | Y |
| thk_left_pericalcarinedktcortex | 0.42 | Y |
| thk_inferior_longitudinal_fasciculus_lwmtracts_left | 0.42 | Y |
| thk_left_precuneusdktregions | 0.42 | Y |
| vol_left_ca1mtl | 0.42 | Y |
| thk_left_pars_opercularisdktregions | 0.42 | Y |
| vol_right_caudatedktregions | 0.42 | Y |
| area_frontal_ldktlobes | 0.42 | Y |
| vol_left_entorhinaldktcortex | 0.42 | Y |
| area_left_pars_opercularisdktregions | 0.42 | Y |
| thk_occipital_rdktlobes | 0.42 | Y |
| vol_occipital_ldktlobes | 0.42 | Y |
| area_corticospinal_lwmtracts_left | 0.42 | Y |
| area_right_amygdaladktregions | 0.42 | Y |
| vol_frontal_rdktlobes | 0.42 | Y |
| thk_left_entorhinaldktcortex | 0.42 | Y |
| vol_die_sth_rightcit168 | 0.42 | Y |
| vol_mtg_rn_leftcit168 | 0.42 | Y |
| thk_inferior_fronto_occipital_rwmtracts_right | 0.42 | Y |
| vol_left_superior_temporaldktregions | 0.42 | Y |
| thk_right_fusiformdktcortex | 0.42 | Y |
| area_left_inferior_parietaldktcortex | 0.42 | Y |
| vol_right_posterior_cingulatedktregions | 0.42 | Y |
| area_left_insuladktregions | 0.42 | Y |

| | | |
|---|---|---|
| area_right_entorhinaldktcortex | 0.42 | Y |
| area_parietal_ldktlobes | 0.42 | Y |
| thk_right_insuladktcortex | 0.42 | Y |
| thk_mtg_rn_leftdeep_cit168 | 0.42 | Y |
| area_left_superior_temporaldktregions | 0.42 | Y |
| thk_left_pars_orbitalisdktregions | 0.42 | Y |
| area_thm_eth_hn_leftcit168 | 0.42 | Y |
| area_frontal_rdktlobes | 0.42 | Y |
| vol_left_fusiformdktregions | 0.42 | Y |
| thk_mtg_sn_snr_leftsnseg | 0.42 | Y |
| area_bn_str_nac_leftcit168 | 0.42 | Y |
| thk_right_medial_orbitofrontaldktcortex | 0.42 | Y |
| thk_left_perirhinalmtl | 0.42 | Y |
| area_right_rostral_anterior_cingulatedktcortex | 0.42 | Y |
| vol_right_cuneusdktregions | 0.42 | Y |
| thk_right_inferior_temporaldktcortex | 0.42 | Y |
| vol_mtg_sn_snc_rightdeep_cit168 | 0.42 | Y |
| thk_right_lateral_occipitaldktcortex | 0.42 | Y |
| area_right_insuladktcortex | 0.42 | Y |
| vol_left_lateral_ventricledktregions | 0.42 | Y |
| vol_inferior_fronto_occipital_lwmtracts_left | 0.42 | Y |
| thk_die_sth_leftcit168 | 0.42 | Y |
| thk_right_isthmus_cingulatedktcortex | 0.42 | Y |
| area_right_parahippocampaldktregions | 0.42 | Y |
| vol_temporal_ldktlobes | 0.42 | Y |
| vol_left_supramarginaldktcortex | 0.42 | Y |
| thk_left_cerebellem_exteriordktregions | 0.42 | Y |
| vol_left_subiculummtl | 0.42 | Y |
| thk_exa_leftcit168 | 0.42 | Y |
| area_csftissues | 0.42 | Y |
| area_mtg_sn_snc_leftcit168 | 0.42 | Y |
| thk_right_pars_orbitalisdktcortex | 0.42 | Y |
| thk_right_entorhinaldktregions | 0.42 | Y |
| vol_right_lateral_orbitofrontaldktcortex | 0.42 | Y |
| vol_left_insuladktcortex | 0.42 | Y |
| area_left_transverse_temporaldktcortex | 0.42 | Y |
| area_right_pericalcarinedktcortex | 0.42 | Y |
| area_right_postcentraldktregions | 0.42 | Y |
| area_occipital_ldktlobes | 0.42 | Y |

| | | |
|---|---|---|
| vol_cerebellar_vermal_lobules_i.vdktregions | 0.42 | Y |
| vol_corpus_callosumwmtracts_right | 0.42 | Y |
| area_right_putamendktregions | 0.42 | Y |
| area_right_perirhinalmtl | 0.42 | Y |
| vol_left_posterior_cingulatedktregions | 0.42 | Y |
| vol_left_cerebellum_white_matterdktregions | 0.42 | Y |
| thk_exa_rightcit168 | 0.42 | Y |
| vol_left_postcentraldktcortex | 0.42 | Y |
| vol_brainstem_ldktlobes | 0.42 | Y |
| thk_right_pars_triangularisdktcortex | 0.42 | Y |
| vol_left_inferior_parietaldktregions | 0.42 | Y |
| area_right_parahippocampaldktcortex | 0.42 | Y |
| thk_left_entorhinaldktregions | 0.42 | Y |
| vol_thm_eth_hn_leftcit168 | 0.42 | Y |
| vol_left_parahippocampaldktcortex | 0.42 | Y |
| thk_die_hth_leftcit168 | 0.42 | Y |
| vol_gmtissues | 0.42 | Y |
| thk_left_supramarginaldktregions | 0.42 | Y |
| vol_right_lingualdktcortex | 0.42 | Y |
| vol_mtg_sn_snr_rightcit168 | 0.42 | Y |
| vol_bn_gp_gpi_leftdeep_cit168 | 0.42 | Y |
| thk_right_caudal_middle_frontaldktregions | 0.42 | Y |
| area_right_medial_orbitofrontaldktcortex | 0.42 | Y |
| area_occipital_rdktlobes | 0.42 | Y |
| thk_left_lateral_occipitaldktregions | 0.42 | Y |
| vol_left_superior_frontaldktcortex | 0.42 | Y |
| vol_left_pars_triangularisdktcortex | 0.42 | Y |
| vol_left_cuneusdktcortex | 0.42 | Y |
| area_corticospinal_rwmtracts_right | 0.42 | Y |
| vol_mtg_sn_snr_rightdeep_cit168 | 0.42 | Y |
| thk_left_supramarginaldktcortex | 0.42 | Y |
| area_left_pars_triangularisdktcortex | 0.42 | Y |
| thk_left_pericalcarinedktregions | 0.42 | Y |
| thk_parietal_ldktlobes | 0.42 | Y |
| vol_left_medial_orbitofrontaldktcortex | 0.42 | Y |
| area_right_superior_frontaldktregions | 0.42 | Y |
| thk_csftissues | 0.42 | Y |
| thk_right_postcentraldktregions | 0.42 | Y |
| vol_left_medial_orbitofrontaldktregions | 0.42 | Y |

| | | |
|---|---|---|
| vol_right_amygdaladktregions | 0.42 | Y |
| thk_right_parahippocampalmtl | 0.42 | Y |
| thk_mtg_sn_snc_leftdeep_cit168 | 0.42 | Y |
| vol_left_entorhinaldktregions | 0.42 | Y |
| vol_left_middle_temporaldktregions | 0.42 | Y |
| thk_right_posterior_cingulatedktregions | 0.42 | Y |
| vol_right_hippocampushipplr | 0.42 | Y |
| area_right_cuneusdktregions | 0.42 | Y |
| vol_mtg_sn_snr_rightsnseg | 0.42 | Y |
| thk_nbm_left_midbf | 0.42 | Y |
| thk_right_lateral_occipitaldktregions | 0.42 | Y |
| vol_left_pars_orbitalisdktregions | 0.42 | Y |
| vol_left_postcentraldktregions | 0.42 | Y |
| area_left_rostral_anterior_cingulatedktcortex | 0.42 | Y |
| thk_right_precentraldktcortex | 0.42 | Y |
| area_right_ca1mtl | 0.42 | Y |
| vol_left_pars_orbitalisdktcortex | 0.42 | Y |
| vol_right_pars_orbitalisdktregions | 0.42 | Y |
| vol_right_lateral_occipitaldktregions | 0.42 | Y |
| vol_mtg_rn_rightcit168 | 0.42 | Y |
| vol_left_caudal_middle_frontaldktcortex | 0.42 | Y |
| vol_right_transverse_temporaldktcortex | 0.42 | Y |
| thk_bn_str_pu_rightcit168 | 0.42 | Y |
| area_left_inferior_temporaldktcortex | 0.42 | Y |
| area_left_pars_orbitalisdktcortex | 0.42 | Y |
| thk_bn_gp_gpi_rightdeep_cit168 | 0.42 | Y |
| vol_right_thalamus_properdktregions | 0.42 | Y |
| area_left_caudal_middle_frontaldktregions | 0.42 | Y |
| area_left_superior_parietaldktcortex | 0.42 | Y |
| vol_parietal_rdktlobes | 0.42 | Y |
| thk_left_rostral_anterior_cingulatedktregions | 0.42 | Y |
| vol_left_posterior_cingulatedktcortex | 0.42 | Y |
| thk_right_inferior_temporaldktregions | 0.42 | Y |
| vol_mtg_vtr_vta_rightcit168 | 0.42 | Y |
| thk_bn_str_nac_rightcit168 | 0.42 | Y |
| area_left_supramarginaldktcortex | 0.42 | Y |
| area_inferior_fronto_occipital_rwmtracts_right | 0.42 | Y |
| vol_right_caudal_anterior_cingulatedktregions | 0.42 | Y |
| vol_right_superior_parietaldktcortex | 0.42 | Y |

| | | |
|---|---|---|
| area_referenceregion_leftdeep_cit168 | 0.42 | Y |
| area_right_superior_temporaldktregions | 0.42 | Y |
| thk_right_paracentraldktregions | 0.42 | Y |
| thk_left_amygdaladktregions | 0.42 | Y |
| thk_right_superior_temporaldktcortex | 0.42 | Y |
| vol_right_precentraldktregions | 0.42 | Y |
| thk_referenceregion_rightcit168 | 0.42 | Y |
| thk_right_hippocampushipplr | 0.42 | Y |
| thk_left_superior_frontaldktcortex | 0.42 | Y |
| thk_bn_gp_gpe_rightcit168 | 0.42 | Y |
| vol_cerebellum_ldktlobes | 0.42 | Y |
| area_bn_str_pu_rightdeep_cit168 | 0.42 | Y |
| thk_brainstemtissues | 0.42 | Y |
| thk_right_rostral_middle_frontaldktregions | 0.42 | Y |
| thk_right_medial_orbitofrontaldktregions | 0.42 | Y |
| area_right_transverse_temporaldktregions | 0.42 | Y |
| vol_left_transverse_temporaldktcortex | 0.42 | Y |
| thk_bn_str_pu_rightdeep_cit168 | 0.42 | Y |
| area_left_parahippocampaldktregions | 0.42 | Y |
| thk_right_transverse_temporaldktregions | 0.42 | Y |
| area_mtg_sn_snr_rightcit168 | 0.42 | Y |
| thk_temporal_ldktlobes | 0.42 | Y |
| thk_left_transverse_temporaldktregions | 0.42 | Y |
| thk_mtg_sn_snr_rightsnseg | 0.42 | Y |
| vol_bn_gp_gpe_rightdeep_cit168 | 0.42 | Y |
| area_left_isthmus_cingulatedktcortex | 0.42 | Y |
| area_left_inferior_parietaldktregions | 0.42 | Y |
| vol_left_caudal_anterior_cingulatedktcortex | 0.42 | Y |
| vol_right_parahippocampaldktregions | 0.42 | Y |
| area_left_fusiformdktregions | 0.42 | Y |
| thk_left_caudal_middle_frontaldktregions | 0.42 | Y |
| vol_right_cerebellum_white_matterdktregions | 0.42 | Y |
| vol_left_putamendktregions | 0.42 | Y |
| vol_left_lateral_orbitofrontaldktcortex | 0.42 | Y |
| vol_left_pericalcarinedktcortex | 0.42 | Y |
| vol_die_hth_mn_rightcit168 | 0.42 | Y |
| vol_left_superior_frontaldktregions | 0.42 | Y |
| area_left_caudal_middle_frontaldktcortex | 0.42 | Y |
| area_right_inferior_parietaldktcortex | 0.42 | Y |

| | | |
|---|---|---|
| area_mtg_rn_leftdeep_cit168 | 0.42 | Y |
| area_left_middle_temporaldktcortex | 0.42 | Y |
| vol_left_rostral_anterior_cingulatedktregions | 0.42 | Y |
| vol_left_superior_parietaldktregions | 0.42 | Y |
| thk_right_lateral_ventricledktregions | 0.42 | Y |
| vol_right_paracentraldktregions | 0.42 | Y |
| area_mtg_sn_snr_rightsnseg | 0.42 | Y |
| vol_right_medial_orbitofrontaldktcortex | 0.42 | Y |
| area_left_precuneusdktcortex | 0.42 | Y |
| thk_left_lateral_orbitofrontaldktcortex | 0.42 | Y |
| area_mtg_sn_snc_leftdeep_cit168 | 0.42 | Y |
| thk_right_insuladktregions | 0.42 | Y |
| area_right_rostral_middle_frontaldktcortex | 0.42 | Y |
| thk_right_palladiumdktregions | 0.42 | Y |
| vol_right_inferior_temporaldktcortex | 0.42 | Y |
| area_mtg_sn_snr_leftdeep_cit168 | 0.42 | Y |
| area_nbm_left_midbf | 0.42 | Y |
| vol_mtg_sn_snc_leftdeep_cit168 | 0.42 | Y |
| vol_left_inferior_temporaldktregions | 0.42 | Y |
| area_right_middle_temporaldktcortex | 0.42 | Y |
| area_left_hippocampusdktregions | 0.42 | Y |
| thk_left_superior_temporaldktcortex | 0.42 | Y |
| vol_left_perirhinalmtl | 0.42 | Y |
| vol_left_lateral_occipitaldktcortex | 0.42 | Y |
| vol_right_superior_temporaldktregions | 0.42 | Y |
| thk_die_sth_rightcit168 | 0.42 | Y |
| vol_mtg_sn_snc_leftcit168 | 0.42 | Y |
| thk_left_pars_orbitalisdktcortex | 0.42 | Y |
| vol_right_cerebellum_exteriordktregions | 0.42 | Y |
| vol_bn_str_pu_rightdeep_cit168 | 0.42 | Y |
| area_parietal_rdktlobes | 0.42 | Y |
| vol_right_fusiformdktcortex | 0.42 | Y |
| area_right_inferior_parietaldktregions | 0.42 | Y |
| area_right_pars_orbitalisdktcortex | 0.42 | Y |
| vol_bn_str_pu_leftcit168 | 0.42 | Y |
| vol_left_parahippocampalmtl | 0.42 | Y |
| area_right_inferior_temporaldktcortex | 0.42 | Y |
| vol_right_subiculummtl | 0.42 | Y |
| area_right_lingualdktregions | 0.42 | Y |

| | | |
|---|---|---|
| area_right_entorhinaldktregions | 0.42 | Y |
| area_right_transverse_temporaldktcortex | 0.42 | Y |
| area_referenceregion_leftcit168 | 0.42 | Y |
| thk_frontal_rdktlobes | 0.42 | Y |
| area_die_sth_leftcit168 | 0.42 | Y |
| vol_parietal_ldktlobes | 0.42 | Y |
| area_left_superior_frontaldktregions | 0.42 | Y |
| thk_right_fusiformdktregions | 0.41 | Y |
| area_superior_longitudinal_fasciculus_lwmtracts_left | 0.41 | Y |
| area_cerebellar_vermal_lobules_viii.xdktregions | 0.41 | Y |
| thk_temporal_rdktlobes | 0.41 | Y |
| area_bn_str_ca_leftcit168 | 0.41 | Y |
| thk_left_superior_parietaldktcortex | 0.41 | Y |
| area_uncinate_rwmtracts_right | 0.41 | Y |
| thk_left_postcentraldktregions | 0.41 | Y |
| vol_brain_stemdktregions | 0.41 | Y |
| thk_right_lateral_orbitofrontaldktcortex | 0.41 | Y |
| area_brainstem_rdktlobes | 0.41 | Y |
| area_cerebellumtissues | 0.41 | Y |
| thk_left_cuneusdktregions | 0.41 | Y |
| vol_right_paracentraldktcortex | 0.41 | Y |
| area_left_lateral_orbitofrontaldktregions | 0.41 | Y |
| area_left_paracentraldktregions | 0.41 | Y |
| area_bn_gp_gpi_rightcit168 | 0.41 | Y |
| area_left_medial_orbitofrontaldktcortex | 0.41 | Y |
| vol_left_superior_parietaldktcortex | 0.41 | Y |
| area_right_posterior_cingulatedktregions | 0.41 | Y |
| area_mtg_rn_rightcit168 | 0.41 | Y |
| thk_left_insuladktcortex | 0.41 | Y |
| vol_nbm_right_midbf | 0.41 | Y |
| vol_right_palladiumdktregions | 0.41 | Y |
| area_mtg_sn_snc_rightcit168 | 0.41 | Y |
| area_right_cuneusdktcortex | 0.41 | Y |
| vol_right_superior_frontaldktcortex | 0.41 | Y |
| vol_right_supramarginaldktcortex | 0.41 | Y |
| thk_left_rostral_middle_frontaldktregions | 0.41 | Y |
| vol_mtg_sn_snr_leftsnseg | 0.41 | Y |
| thk_left_hippocampusdktregions | 0.41 | Y |
| thk_die_hth_mn_rightcit168 | 0.41 | Y |

| | | |
|---|---|---|
| vol_left_caudatedktregions | 0.41 | Y |
| thk_left_lateral_ventricledktregions | 0.41 | Y |
| area_right_precuneusdktcortex | 0.41 | Y |
| thk_cerebellum_rdktlobes | 0.41 | Y |
| vol_left_ventral_dcdktregions | 0.41 | Y |
| thk_left_lingualdktcortex | 0.41 | Y |
| thk_right_pericalcarinedktregions | 0.41 | Y |
| area_right_lateral_orbitofrontaldktcortex | 0.41 | Y |
| thk_right_hippocampusdktregions | 0.41 | Y |
| thk_left_superior_frontaldktregions | 0.41 | Y |
| vol_left_lingualdktregions | 0.41 | Y |
| vol_mtg_rn_leftdeep_cit168 | 0.41 | Y |
| thk_4th_ventricledktregions | 0.41 | Y |
| thk_right_middle_temporaldktcortex | 0.41 | Y |
| vol_right_ventral_dcdktregions | 0.41 | Y |
| area_bn_str_ca_rightcit168 | 0.41 | Y |
| vol_referenceregion_leftcit168 | 0.41 | Y |
| thk_left_precentraldktcortex | 0.41 | Y |
| vol_right_superior_frontaldktregions | 0.41 | Y |
| vol_inferior_longitudinal_fasciculus_lwmtracts_left | 0.41 | Y |
| thk_right_paracentraldktcortex | 0.41 | Y |
| area_right_superior_parietaldktregions | 0.41 | Y |
| vol_left_amygdaladktregions | 0.41 | Y |
| wmh_integral_prob | 0.41 | Y |
| area_wmtissues | 0.41 | Y |
| vol_left_transverse_temporaldktregions | 0.41 | Y |
| thk_brain_stemdktregions | 0.41 | Y |
| vol_right_middle_temporaldktregions | 0.41 | Y |
| area_left_ventral_dcdktregions | 0.41 | Y |
| area_left_cuneusdktregions | 0.41 | Y |
| area_right_alecmtl | 0.41 | Y |
| thk_right_caudal_anterior_cingulatedktcortex | 0.41 | Y |
| area_referenceregion_rightcit168 | 0.41 | Y |
| area_right_caudal_anterior_cingulatedktregions | 0.41 | Y |
| vol_right_lateral_ventricledktregions | 0.41 | Y |
| area_cerebellum_ldktlobes | 0.41 | Y |
| vol_right_rostral_anterior_cingulatedktcortex | 0.41 | Y |
| thk_mtg_sn_snc_rightcit168 | 0.41 | Y |
| vol_right_posterior_cingulatedktcortex | 0.41 | Y |

| | | |
|---|---|---|
| thk_die_hth_rightcit168 | 0.41 | Y |
| thk_left_paracentraldktregions | 0.41 | Y |
| vol_right_rostral_middle_frontaldktcortex | 0.41 | Y |
| vol_deepgraytissues | 0.41 | Y |
| thk_right_superior_frontaldktcortex | 0.41 | Y |
| area_right_caudatedktregions | 0.41 | Y |
| area_left_paracentraldktcortex | 0.41 | Y |
| thk_left_caudal_anterior_cingulatedktcortex | 0.41 | Y |
| area_brainstem_ldktlobes | 0.41 | Y |
| thk_superior_longitudinal_fasciculus_rwmtracts_right | 0.41 | Y |
| vol_right_hippocampusdktregions | 0.41 | Y |
| vol_left_hippocampushipplr | 0.41 | Y |
| vol_left_caudal_anterior_cingulatedktregions | 0.41 | Y |
| vol_right_superior_parietaldktregions | 0.41 | Y |
| vol_uncinate_rwmtracts_right | 0.41 | Y |
| area_right_medial_orbitofrontaldktregions | 0.41 | Y |
| vol_bn_gp_gpi_leftcit168 | 0.41 | Y |
| thk_thm_eth_hn_rightcit168 | 0.41 | Y |
| vol_cerebellum_rdktlobes | 0.41 | Y |
| area_mtg_sn_snr_leftcit168 | 0.41 | Y |
| vol_inferior_fronto_occipital_rwmtracts_right | 0.41 | Y |
| area_left_postcentraldktcortex | 0.41 | Y |
| area_right_cerebellum_white_matterdktregions | 0.41 | Y |
| thk_left_rostral_middle_frontaldktcortex | 0.41 | Y |
| vol_left_precentraldktcortex | 0.41 | Y |
| thk_mtg_sn_snr_rightdeep_cit168 | 0.41 | Y |
| vol_left_rostral_anterior_cingulatedktcortex | 0.41 | Y |
| area_right_caudal_middle_frontaldktregions | 0.41 | Y |
| thk_left_inferior_temporaldktcortex | 0.41 | Y |
| vol_superior_longitudinal_fasciculus_lwmtracts_left | 0.41 | Y |
| area_left_parahippocampaldktcortex | 0.41 | Y |
| thk_right_cuneusdktregions | 0.41 | Y |
| thk_right_ventral_dcdktregions | 0.41 | Y |
| area_left_rostral_middle_frontaldktcortex | 0.41 | Y |
| area_right_paracentraldktregions | 0.4 | Y |
| area_mtg_vtr_pbp_leftcit168 | 0.4 | Y |
| vol_left_cuneusdktregions | 0.4 | Y |
| area_left_caudatedktregions | 0.4 | Y |
| area_right_superior_temporaldktcortex | 0.4 | Y |

| | | |
|---|---|---|
| vol_left_cerebellem_exteriordktregions | 0.4 | Y |
| area_gmtissues | 0.4 | Y |
| area_bn_gp_gpe_leftcit168 | 0.4 | Y |
| vol_right_entorhinaldktregions | 0.4 | Y |
| area_left_transverse_temporaldktregions | 0.4 | Y |
| thk_left_ca1mtl | 0.4 | Y |
| area_mtg_rn_leftcit168 | 0.4 | Y |
| area_right_rostral_anterior_cingulatedktregions | 0.4 | Y |
| thk_left_pallidiumdktregions | 0.4 | Y |
| thk_right_supramarginaldktregions | 0.4 | Y |
| vol_bn_str_pu_rightcit168 | 0.4 | Y |
| thk_left_posterior_cingulatedktregions | 0.4 | Y |
| vol_right_caudal_middle_frontaldktregions | 0.4 | Y |
| vol_left_rostral_middle_frontaldktcortex | 0.4 | Y |
| thk_referenceregion_leftcit168 | 0.4 | Y |
| vol_right_superior_temporaldktcortex | 0.4 | Y |
| vol_left_pallidiumdktregions | 0.4 | Y |
| thk_right_inferior_parietaldktcortex | 0.4 | Y |
| vol_right_inferior_temporaldktregions | 0.4 | Y |
| thk_right_rostral_anterior_cingulatedktregions | 0.4 | Y |
| area_left_alecmtl | 0.4 | Y |
| area_cerebellar_vermal_lobules_vi.viidktregions | 0.4 | Y |
| vol_right_precentraldktcortex | 0.4 | Y |
| vol_right_inferior_parietaldktcortex | 0.4 | Y |
| vol_right_inferior_parietaldktregions | 0.4 | Y |
| vol_left_isthmus_cingulatedktregions | 0.4 | Y |
| area_right_palladiumdktregions | 0.4 | Y |
| area_right_supramarginaldktcortex | 0.4 | Y |
| area_left_posterior_cingulatedktregions | 0.4 | Y |
| area_right_isthmus_cingulatedktregions | 0.4 | Y |
| area_left_lateral_occipitaldktcortex | 0.39 | Y |
| area_right_hippocampusdktregions | 0.39 | Y |
| thk_right_parahippocampaldktcortex | 0.39 | Y |
| vol_right_rostral_middle_frontaldktregions | 0.39 | Y |
| brainVolume | 0.34 | Y |
| RandBasisProjPos10 | 0.25 | Y |
| RandBasisProjPos08 | 0.25 | Y |
| RandBasisProj07 | 0.25 | Y |
| RandBasisProjPos02 | 0.25 | Y |

| | | |
|---|---|---|
| RandBasisProj03 | 0.25 | Y |
| RandBasisProj05 | 0.25 | Y |
| RandBasisProjPos07 | 0.25 | Y |
| RandBasisProj01 | 0.25 | Y |
| RandBasisProj04 | 0.25 | Y |
| RandBasisProjPos06 | 0.25 | Y |
| RandBasisProj08 | 0.25 | Y |
| RandBasisProj06 | 0.25 | Y |
| RandBasisProjPos09 | 0.25 | Y |
| RandBasisProjPos01 | 0.25 | Y |
| RandBasisProj09 | 0.25 | Y |
| RandBasisProjPos03 | 0.24 | Y |
| RandBasisProj10 | 0.24 | Y |
| RandBasisProj02 | 0.24 | Y |
| RandBasisProjPos05 | 0.24 | Y |
| RandBasisProjPos04 | 0.24 | Y |

**Supplementary Table 2. Top hyperparameters for trained models and ensemble weights.**

| Task | Model name | Top hyperparameter | Ensemble weight |
|---|---|---|---|
| AD prediction task | Logistic Regression | {'solver': 'saga', 'max_iter': 100, 'C': 275.25, 'penalty': 'l1'} | 0.44 |
| | Neural Network | {'FeatureSelection': 'LASSO', 'optimizer': 'adamw', 'random_state': 42, 'layers': 4, 'layer_size0': 135, 'layer_size1': 275, 'layer_size2': 141, 'layer_size3': 11, 'dropout': 0.17, 'learning_rate': 0.0001, 'weight_decay': 0.0, 'lasso_c': 0.06} | 0.50 |
| | XGB Classifier | {'n_estimators': 15, 'booster': 'gbtree', 'learning_rate': 0.09, 'reg_lambda': 1.89, 'gamma': 0.0, 'reg_alpha': 0.02, 'max_depth': 4, 'min_child_weight': 0.01, 'subsample': 0.4, 'colsample_bytree': 0.17, 'tree_method': 'gpu_hist', 'predictor': 'cpu_predictor', 'max_bin': 80, 'FeatureSelection': 'LASSO', 'colsample_bylevel': 0.88, 'early_stopping_rounds': 75, 'max_leaves': 80, 'scale_pos_weight': 2.72, 'lasso_c': 8.07} | 0.06 |
| PD prediction task | Logistic Regression | {'solver': 'saga', 'max_iter': 100, 'C': 184.46, 'penalty': 'l2'} | 0.42 |

| | | | |
|---|---|---|---|
| | Neural Network | {'max_epochs': 500, 'optimizer': 'adamw', 'early_stopping': 1, 'random_state': 42, 'layers': 2, 'layer_size0': 85, 'layer_size1': 337, 'dropout': 0.46, 'learning_rate': 0.0001, 'weight_decay': 0.0} | 0.55 |
| | XGB Classifier | {'n_estimators': 45, 'booster': 'gbtree', 'learning_rate': 0.0, 'reg_lambda': 7.55, 'gamma': 40.45, 'reg_alpha': 0.0, 'max_depth': 10, 'min_child_weight': 0.0, 'subsample': 0.52, 'colsample_bytree': 0.69, 'tree_method': 'gpu_hist', 'predictor': 'cpu_predictor', 'max_bin': 289, 'FeatureSelection': 'LASSO', 'colsample_bylevel': 0.1, 'early_stopping_rounds': 45, 'max_leaves': 740, 'scale_pos_weight': 3.54} | 0.03 |
